# Windows of Susceptibility to Air Pollution During and Surrounding Pregnancy in Relation to Longitudinal Maternal Measures of Adiposity and Lipid Profiles

**DOI:** 10.1101/2024.11.23.24317830

**Authors:** Sandra India Aldana, Danielle Demateis, Damaskini Valvi, Allan C. Just, Iván Gutiérrez-Avila, Guadalupe Estrada-Gutierrez, Martha María Téllez Rojo, Robert O. Wright, Andrea A. Baccarelli, Haotian Wu, Kayleigh P. Keller, Ander Wilson, Elena Colicino

**Author notes:** Corresponding author: Sandra India Aldana, MPH, MPhil, PhD Postdoctoral Fellow Department of Environmental Medicine and Climate Science Icahn School of Medicine at Mount Sinai 17 E 102 St, Floor 3 New York, NY 10029.

## Abstract

Pregnancy is a critical window for long-term metabolic programming of fetal effects stemming from airborne particulate matter ≤2.5μm (PM_2.5_) exposure. Yet, little is known about long-term metabolic effects of PM_2.5_ exposure during and surrounding pregnancy in mothers. We assessed potential critical windows of PM_2.5_ exposure during and surrounding pregnancy with maternal adiposity and lipid measures later in life. We included 517 pregnant women from the PROGRESS cohort with adiposity [body mass index (BMI), waist circumference (WC), % body fat] and lipids [total cholesterol, high-density-lipoprotein (HDL), low-density-lipoprotein (LDL)] measured repeatedly at 4, 6 and 8 years post-delivery. Monthly average PM_2.5_ exposure was estimated at each participant’s address using a validated spatiotemporal model. We employed distributed lag interaction models (DLIMs) adjusting for socio-demographics and clinical covariates. We found that a 1 μg/m^3^ increase in PM_2.5_ exposure throughout mid-/late-pregnancy was associated with higher WC at 6-years post-delivery, peaking at 6 months of gestation: 0.04 cm (95%CI: 0.01, 0.06). We also identified critical windows of PM_2.5_ exposure during and surrounding pregnancy associated with higher LDL and lower HDL both measured at 4 years post-delivery with peaks at pre-conception for LDL [0.17 mg/dL (95%CI: 0.00, 0.34)] and at the 11^th^ month after conception for HDL [-0.07 mg/dL (95%CI: -0.11, -0.02)]. Stratified analyses by fetal sex indicated stronger associations with adiposity measures in mothers carrying a male, whereas stronger associations were observed with lipids in mothers carrying a female fetus. Stratified analyses also indicated potential stronger deleterious lagged effects in women with folic acid intake lower than 600mcg/day during pregnancy.

## 1. Introduction

Obesity and dyslipidemia are key cardiometabolic risk factors and conditions that predispose individuals to increased risk of stroke, disability, and mortality.^1,2^ These conditions have increased in North America at an alarming rate throughout the past decades^3–10^ and are particularly more prevalent among Hispanics.^11,12^ In Mexican adults, dyslipidemias are the most prevalent risk factor for cardiovascular diseases,^13^ including hypoalphalipoproteinemia, also known as low high-density lipoprotein (HDL), and high low-density lipoprotein (LDL), with an estimated prevalence of ∼55%. Excess adiposity and low HDL predispose individuals to increased metabolic syndrome, a condition on the rise in Mexico, where it is estimated to affect 56% of the adult population.^6^ Importantly, metabolic disease-related morbidities and mortality affect women more than men in Mexico.^10^ According to the 2018-2019 ENSANUT National Health Nutrition Survey there are substantial higher rates of generalized and abdominal obesity in Mexican adult women compared to men,^14^ with a peak prevalence in women of 40.2% for generalized obesity and 88% for abdominal obesity.

Airborne particulate matter with an aerodynamic diameter of 2.5 microns or smaller (PM_2.5_) is a ubiquitous exposure in urban, industrial, and rural environments.^15,16^ PM_2.5_ exposure can have endocrine-disrupting effects^17–20^ and is considered an emerging risk factor for obesity and adverse metabolic outcomes.^21–23^ Experimental studies in animals and epidemiological research in humans have demonstrated that PM_2.5_ exposure may promote weight gain and adipose tissue expansion, and alter lipid metabolism.^24–29^ Mechanisms by which PM_2.5_ could influence metabolic diseases include hormonal alteration, increased inflammation, oxidative stress, and reactive oxygen species-induced endothelial dysfunction.^30–34^ Furthermore, PM_2.5_ exposure during pregnancy has been suggested to be a critical exposure window for a wide variety of health-related outcomes in both birthing-mothers and children.^35–40^ However, most of the literature examining critical PM_2.5_ exposure windows during pregnancy has primarily focused on children’s health, largely missing potential long-term effects in mothers.^41^ Pregnancy constitutes a major period of physiological changes^42^ including pro-inflammatory processes^43–45^ and weight gain.^46,47^ The numerous physiological changes occurring during pregnancy may lead to potential lingering chronic diseases and metabolic effects.^48–52^ Research on pregnancy as a window of susceptibility to environmental exposures in mothers is scarce^41^ and overlook a potential period in which metabolic conditions may worsen or can effectively be prevented or treated.

To date, no study has evaluated how PM_2.5_ exposure during and surrounding gestation may affect maternal obesity and dyslipidemia in the years following delivery. Therefore, we assessed potential critical windows of PM_2.5_ exposure during and surrounding pregnancy with later changes in markers for anthropometry and lipids in the years following delivery in women from the Mexican Programming Research in Obesity, Growth, Environment and Social Stressors (PROGRESS) study. To achieve this, we employ the novel distributed lag models with interaction to identify critical windows of PM_2.5_ exposure for maternal health endpoints repeatedly measured over post-delivery follow-up visits.^53^ Furthermore, given that maternal metabolic health and associated mechanisms are considered sex-dimorphic^54^ and that folic acid (FA) may attenuate adverse health effects from air pollutants,^55^ we also hypothesized potential effect modification by fetal sex and FA.

## 2. Material and Methods

### 2.1 Study Population

The PROGRESS study enrolled 948 mother-children pairs between 2007-2011 that were followed for over a decade.^56^ Healthy pregnant women, affiliated with the Mexican Social Security Institute, were recruited during the 2^nd^ trimester of pregnancy (12-20 gestational weeks). Any pregnant woman, ≥18 years old and living in Mexico City, with no intention of changing address location for at least three years, were eligible for enrollment. Medical exclusion criteria included history of infertility, heart or renal diseases, diabetes, psychosis or use of anti-epilepsy drugs, use of steroids, consumption of one or more alcoholic drinks per day, or drug addiction. All women enrolled in the study agreed to sign a consent form in Spanish. This study was approved by the Institutional Review Boards of the participating institutions: the Icahn School of Medicine at Mount Sinai and the *“*Instituto Nacional de Salud Pública” in Mexico.

### 2.2 Ambient PM_2.5_ Exposure at Participants’ Addresses

Daily PM_2.5_ predictions with a 1×1-km spatial resolution came from our machine-learning models developed for the Mexico City Metropolitan Area. Our models employed extreme gradient boosting (XGBoost) and inverse-distance weighting (IDW), incorporating satellite-based aerosol optical depth data from NASA’s Terra and Aqua satellites, meteorological information, and land-use variables (**Methods S1**).^57^ To limit the daily exposure variability and facilitate the interpretation of long-term maternal health effects from PM_2.5_, we averaged the daily PM_2.5_ exposures into monthly ambient PM_2.5_ exposure concentrations. We then focused on exposure surrounding pregnancy: pre-conception (approximately one and two months prior to the last menstrual period, LMP), during pregnancy (months 0-9 after LMP), and up to one year after childbirth (months 10-22 after LMP). We examine exposure time up to one year beyond birth as we hypothesize that lingering immediate effects from pregnancy or giving birth could make women more susceptible to environmental exposures.

### 2.3 Outcomes

Repeated anthropometric and lipid measures were collected at three follow-up visits 4, 6 and 8 years post-delivery (corresponding to approximately 48, 72, and 96 months after childbirth). Maternal anthropometric measures (weight, height, waist circumference, and body fat) were collected using standardized protocols.^58–60^ We calculated body mass index (BMI) based on measured height (collected using a Seca 206 roller tape) and weight (collected by a trained physician). Medical staff measured body fat using InBody 270 or InBody 370 Body Composition Analyzers during each monitoring visit. WC was measured using a Seca tape (model 201) above the iliac crest.^60^ Total cholesterol and high-density lipoprotein (HDL) cholesterol were measured by enzymatic photometric assays on a Response 910 automated analyzer (DiaSys Diagnostic Systems, Holzheim, Germany) in fasting plasma collected from participants at all post-delivery visits. Low-density lipoprotein (LDL) concentration was derived using the Friedewald equation.^61^

### 2.4 Additional Variables

Information on socio-demographic factors (age, marital status, socio-economic status) was self-reported at recruitment during the 2^nd^ trimester of pregnancy. PROGRESS also collected lifestyle information at baseline and across follow-up visits: smoking, alcohol intake, medications (**Methods S2**), and parity. Self-reported information on multiple pregnancies throughout the follow-up was also monitored and documented in PROGRESS. Pregnancy start date was defined as the LMP, which was used as a proxy of the month of conception in this study. We used a standard physical assessment of gestational age (Capurro method)^62^ when gestational age differed by more than three weeks from that assigned by the LMP (∼4% of participants in the cohort). We estimated pre-pregnancy BMI in accordance to previous published methods using the height at recruitment and the LMP reported weight.^58,63^ We assessed socio-economic status (SES) using the previously validated Mexican Association of Market and Public Opinion Research Agencies (Spanish acronym AMAI) index used in Mexico.^64^ The AMAI index is an individual-level variable that considered a total of 13 factors including the education of the head of household, number of appliances in the household, car ownership, and so forth. The index denotes six categories that were eventually collapsed into three levels of SES: lower (AMAI categories D and E), medium (AMAI categories C and D+), and higher (AMAI categories A, B, and C+). Our SES measures are relative as the cohort was predominantly low SES. Information on the sex of the child and self-reported FA supplementation intake during pregnancy (at trimesters 2 and 3) were also obtained.

### 2.5 Statistical Analyses

A total of n=806 women had monthly PM_2.5_ exposure from two months prior to LMP through month 22 after LMP and no missing information on pre-pregnancy BMI, age, or meteorological season (**Figure S1**). Missing monthly exposure data during the time period of interest (accounting for <0.3% of the total monthly air pollution data in the cohort) were imputed using time series weighted moving averages.^65^ Women with at least one cardiometabolic marker measured at any follow-up visit were eligible for inclusion resulting in an analytic sample size of n=517.

To evaluate the potential critical windows of PM_2.5_ exposure over 25 months in later onset of cardiometabolic conditions (i.e. higher adiposity and cholesterol levels) at the 4, 6, and 8-year post-delivery visits, we implemented distributed lag interaction models (DLIMs).^53^ The DLIM estimates a linear exposure-time-response function that varies continuously by time after delivery and by outcome assessment time (**Methods S3**). We compared three possible models for how the exposure effect varies across outcome time using a bootstrap likelihood ratio test: no modification, linear modification (i.e. linear scaling of the linear exposure effect across outcome times), or complex non-linear modification across outcome times. In all models, we adjusted for a core set of covariates based on their associations with both the exposure and outcomes or based on their clinical relevance as previously identified (**Figure S2**): age at parturition (continuous),^66^ pre-pregnancy BMI (continuous),^67^ socio-economic status (lower, medium, higher),^68^ smoking during pregnancy (any passive or active smoking reported during pregnancy),^69,70^ marital status (single, separated or divorced vs. married or living with a partner),^71,72^ parity at baseline (1^st^ pregnancy, 2^nd^ pregnancy, and 3^rd^ or more pregnancies at recruitment),^73,74^ season (dry-cold/November to February, dry-warm/March and April, rainy/May to October),^75,76^ medications to treat cardiometabolic diseases^77^ (self-report of a drug treatment for abnormal levels of either glucose, triglycerides, high-density lipoprotein (HDL), or blood pressure vs. lack thereof), alcohol intake (yes/no),^78^ follow-up visit (stage), and follow-up time in months squared to allow for a non-linear effect. Given that lifestyle variables such as alcohol intake, medications, and pregnancy history could change over time, we updated the models with information on lifestyle data at each follow-up stage (at 4, 6, and 8 years post-delivery) as time-varying covariates.

We performed sensitivity analyses by fitting DLIMs 1) without pre-pregnancy BMI as covariate, 2) controlling for time-varying 1-year average PM_2.5_ exposure prior to visit, and 3) without time-varying pregnancy as covariate, generating plots that are smoothed to allow for comparison of sensitivity analyses to our main analyses. Additional analyses were also conducted stratifying by important factors during pregnancy based on prior clinical relevance and/or previous research in PROGRESS. Given the shifting sex-steroid hormone concentrations during pregnancy dependent on fetal sex^79,80^ and the protective interaction effects of FA with air pollutant mixtures and metabolic health in PROGRESS women,^81^ we conducted stratified analyses by 1) fetal sex (mothers carrying a male vs. a female fetus) and by 2) FA intake using the average recommended FA supplementation for pregnant women according to clinical guidelines (600 mcg/day).^82^ All statistical analyses were conducted using R software (version 4.3.0) with package *dlim* (version 0.1.0).

## 3. Results

Women were exposed on average to a PM_2.5_ concentration of 22.8 μg/m^3^ (standard deviation (SD): 1.5; interquartile range (IQR): 2.21) across the 25-month span from 2 month pre-conception up to 1-year post-partum (**Table 1**; **Figure S3**). Women were slightly overweight prior to pregnancy (nearly ∼60% were either overweight or obese) with a mean (SD) BMI of 26.5 (4.1) kg/m^2^ and were on average (SD) 29 (5.6) years old at the time of parturition. The majority of women had low socio-economic status (53%), reported previous pregnancies at baseline prior to index pregnancy (61%), were married or lived with a partner at the time of recruitment (81%), and did not smoke nor were exposed to passive (2^nd^-hand) smoke during pregnancy (63%). Participants were followed for nearly a decade after parturition and attended up to three follow-up examination visits at approximately 4 years [mean (min, max): 57 months (48, 80)], 6 years [mean (min, max): 80 months (71, 115)], and 8 years post-delivery [mean (min, max): 115 months (97, 142)].

**Table 1.**
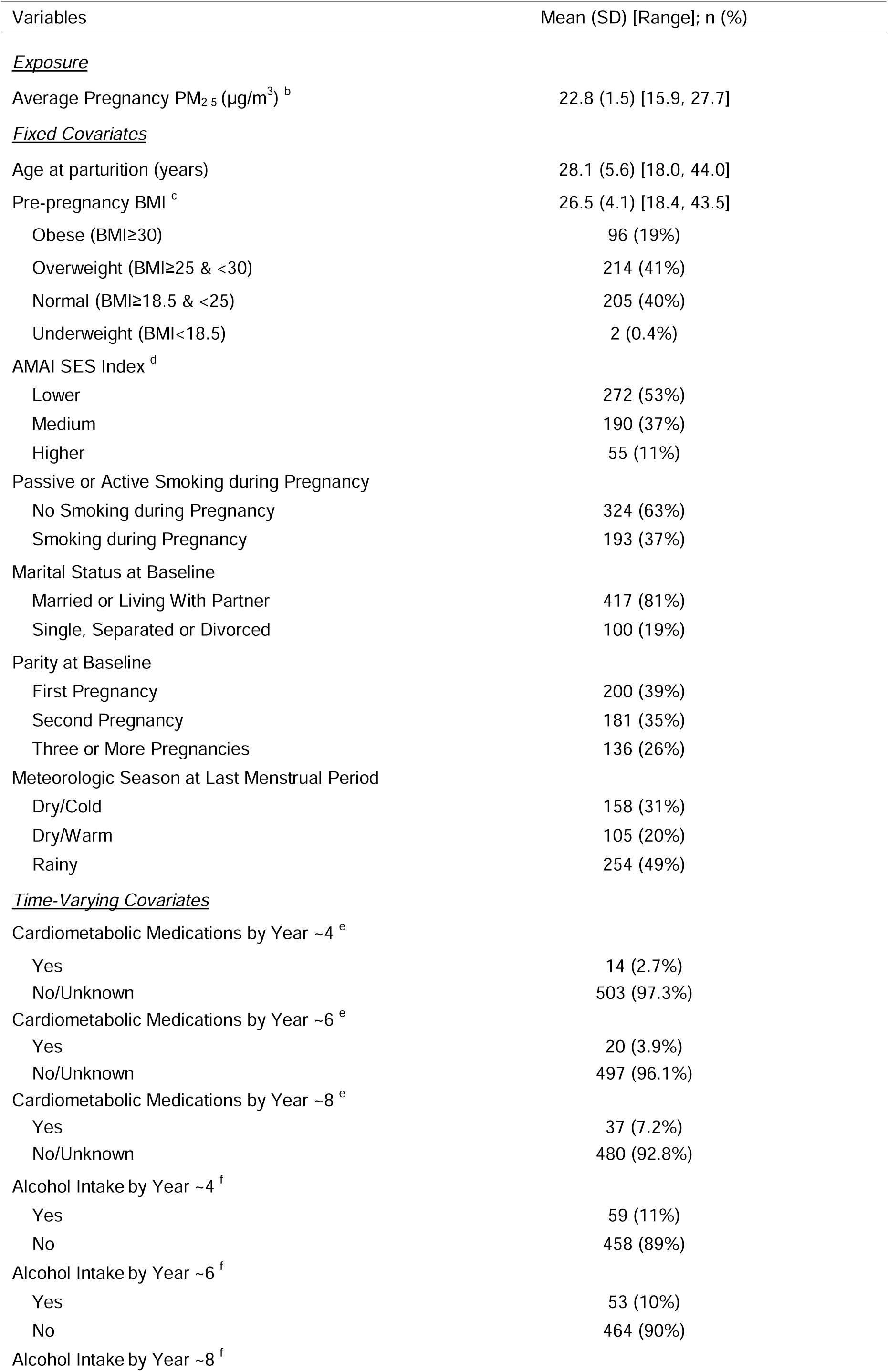
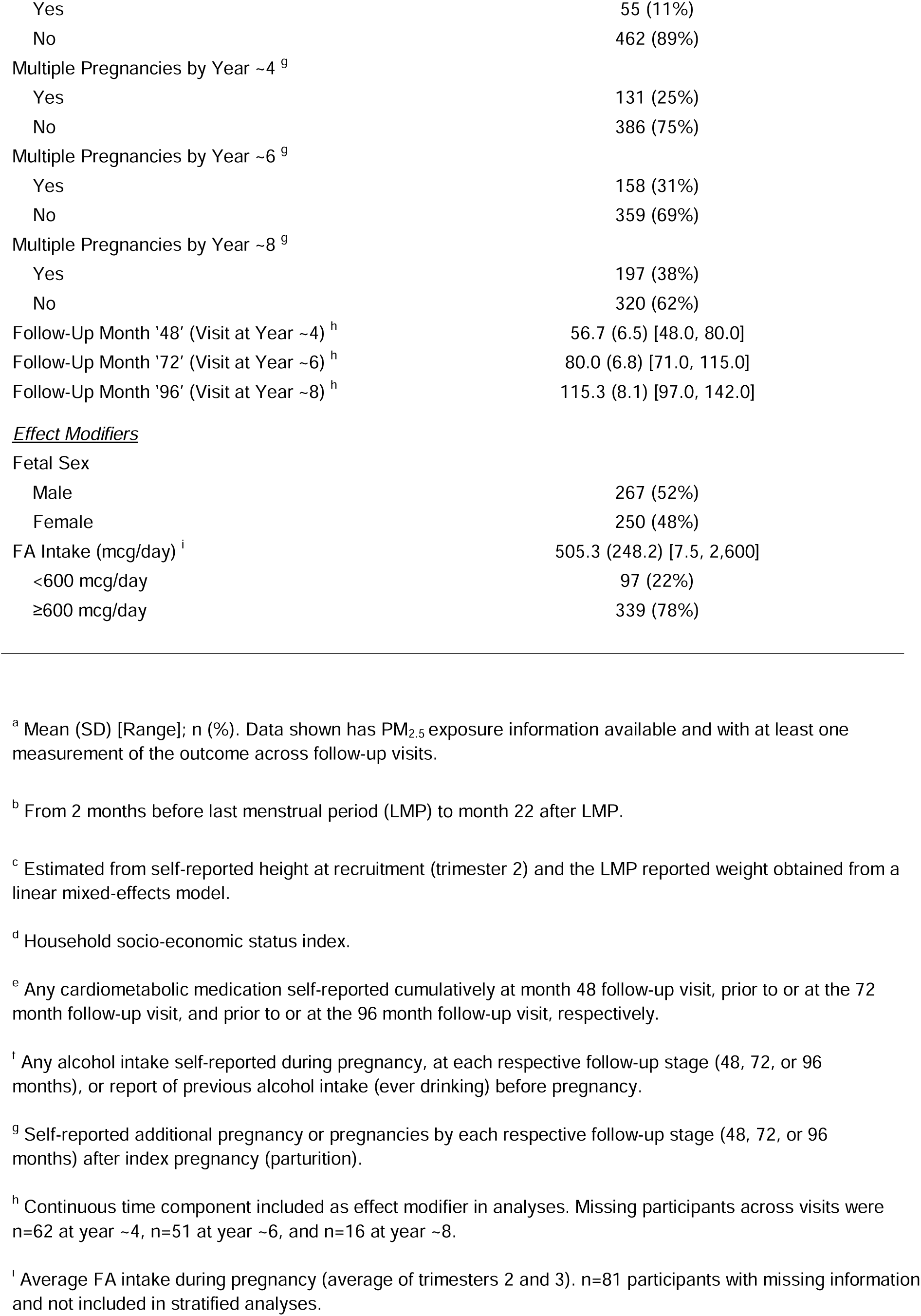
Main Participant Characteristics in PROGRESS (n=517) ^a^.

**Table 2.** Participant Cardiometabolic Outcomes at each Follow-up Visit. ^a^

|  | 48 Months (~4 Years)<br>Mean (SD) [Range] | 72 Months (~6 Years)<br>Mean (SD) [Range] | 96 Months (~8 Years)<br>Mean (SD) [Range] |
| --- | --- | --- | --- |
| *<u>Anthropometric Measures</u>* |  |  |  |
| Body Mass Index (BMI in kg/m<sup>2</sup>) | 27.5 (4.9) [16.4, 45.4]<br>n=455 | 28.0 (5.1) [17.9, 49.6]<br>n=426 | 29.1 (5.2) [16.1, 53.7]<br>n=503 |
| Waist Circumference (WC in cm) | 94.2 (12.2) [63.6, 133.3]<br>n=440 | 92.6 (11.9) [61.1, 132.6]<br>n=453 | 95.3 (13.0) [62.9, 167.6]<br>n=492 |
| Body Fat (%) | 38.1 (6.6) [16.3, 53.9]<br>n=439 | 38.4 (7.0) [3.0, 54.7]<br>n=451 | 40.4 (6.2) [19.9, 54.3]<br>n=493 |
| *<u>Cholesterol Levels</u>* |  |  |  |
| Total Cholesterol (mg/dL) | 185.2 (39.5) [98.0, 441.0]<br>n=447 | 179.8 (32.9) [74.0, 407.0]<br>n=462 | 179.1 (34.1) [91.0, 322.0]<br>n=499 |
| High-Density Lipoprotein (HDL in mg/dL) | 51.3 (12.1) [29.5, 163.0]<br>n=447 | 48.6 (11.9) [10.5, 135.4]<br>n=462 | 47.2 (11.0) [21.0, 93.0]<br>n=499 |
| Low-Density Lipoprotein (LDL in mg/dL) | 108.5 (33.9) [17.3, 308.2]<br>n=447 | 103.6 (27.9) [22.1, 261.6]<br>n=462 | 104.7 (29.0) [10.8, 226.5]<br>n=499 |

<sup>a</sup> Data shown has PM<sub>2.5</sub> exposure information available and was restricted to observations with at least one of the outcomes of interest at each stage.

Using DLIMs, we found evidence of non-linear modification of the exposure-time-response function across outcome times for all outcomes using the bootstrap likelihood ratio test (rejected null of linear modification versus the alternative of non-linear modification). We estimated critical windows of exposure surrounding pregnancy during which PM_2.5_ exposure was linked with several anthropometric measures and lipid levels (**Tables S1-S6**), although these associations were not identified as statistically significant for all cardiometabolic markers examined. Pregnancy PM_2.5_ exposure was associated with changes in several metabolic health measures up to the 6-year post-delivery visit (**Figures 1-2**). A 1-μg/m^3^ increase in PM_2.5_ exposure throughout the 2^nd^ and 3^rd^ trimesters of pregnancy (gestational months: 4-8 after LMP) was associated with an increase in WC levels at the 6-year post-delivery visit (**Figure 1**). The peak effect was at 6 months of gestation which showed an increase of 0.04 cm in WC (95% CI: 0.01, 0.06) per μg/m^3^ PM_2.5_ (**Figure 1**; **Table S2**). A suggestive late pregnancy window of susceptibility to PM_2.5_ exposure was identified in relation to WC levels at the 4-year post-delivery visit [gestational month 9 after LMP: β = 0.03 (95% CI: -0.01, 0.06)]. In addition, we identified alterations in BMI at the 8-year post-delivery visit in association with a peri-conception window of PM_2.5_ exposure (during pre-conception and early-pregnancy period) (**Figure 1**; **Table S1**). These negative PM_2.5_-BMI associations were not sustained in the sensitivity (**Figure S4**) and in the female-stratified (**Figure S8**) analyses, and can be driven by the over-adjustment with pre-pregnancy BMI. There was no evidence of associations in relation to percent body fat.

**Figure 1.**
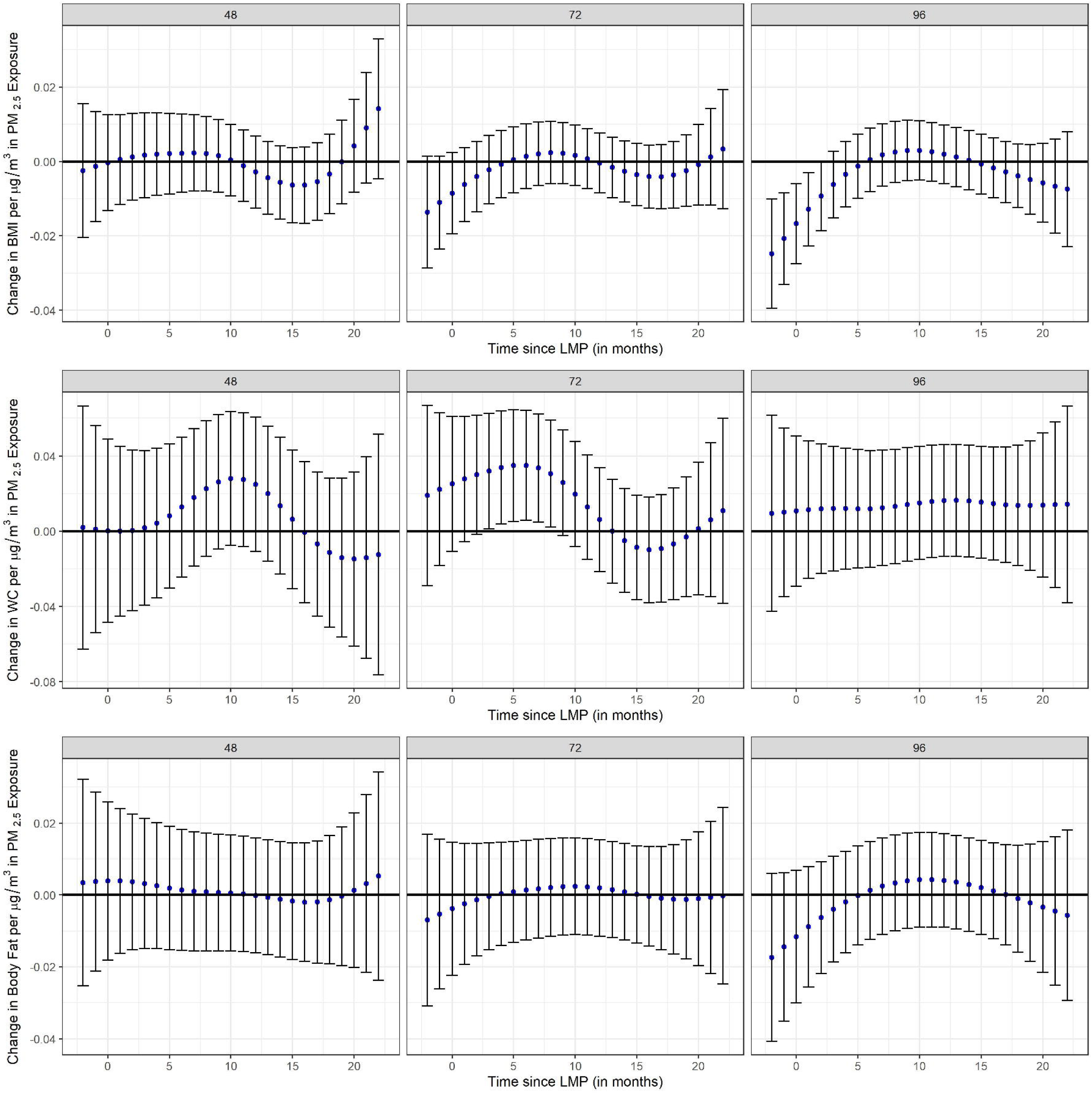
Estimates of the association (blue dots) and 95% confidence interval (95% CI) (grey whiskers) between monthly PM2.5 exposure and anthropometric measures Body Mass Index (BMI), Waist Circumference (WC), body fat percentage at 48, 72, and 96 months (∼4, 6, 8 years) after delivery using distributed lag interaction models (DLIMs) with penalization and non-linear modification. Negative and positive values on the x-axis indicate months before and after last menstrual period (LMP), respectively. Month 0 indicates the LMP month (month 0). Models were adjusted for age, pre-pregnancy BMI, socio-economic status, smoking during pregnancy, marital status, parity at enrollment, meteorological season, cardiometabolic medications, alcohol intake, multiple pregnancies throughout follow-up, stage, and follow-up time in months squared to allow for a non-linear effect. Sample sizes for analyses with BMI were n=516 and sample sizes for analyses with WC and body fat were n=515.

**Figure 2.**
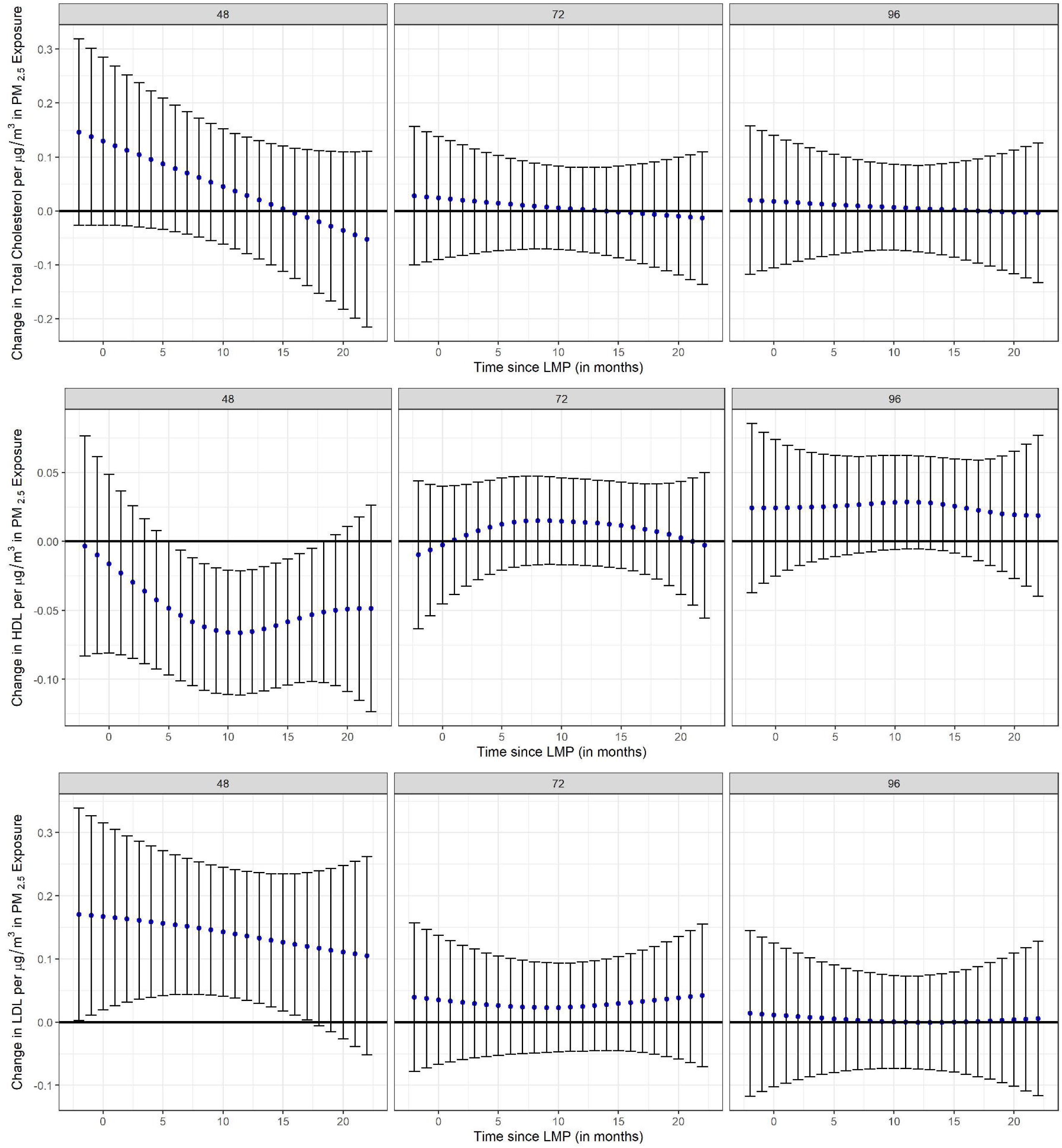
Estimates of the association (blue dots) and 95% confidence interval (95% CI) (grey whiskers) between monthly PM2.5 exposure and total cholesterol, high-density lipoprotein (HDL), low-density lipoprotein (LDL) at 48, 72, and 96 months (∼4, 6, and 8 years) after delivery using distributed lag interaction models (DLIMs) with penalization and non-linear modification. Negative and positive values on the x-axis indicate months before and after last menstrual period (LMP), respectively. Month 0 indicates the LMP month (month 0). Models were adjusted for age, pre-pregnancy BMI, socio-economic status, smoking during pregnancy, marital status, parity at enrollment, meteorological season, cardiometabolic medications, alcohol intake, multiple pregnancies throughout follow-up, stage, and follow-up time in months squared to allow for a nonlinear effect. Sample sizes for analyses with total cholesterol, HDL, and LDL were n=517.

PM_2.5_ exposure surrounding pregnancy was associated with a dysregulation of several lipid levels at the 4-year post-delivery visit, but not thereafter (**Figure 2**). We observed a susceptible PM_2.5_ exposure window from the 2^nd^ trimester of pregnancy and up to the early post-partum period (critical window between months 6 and 18 after LMP) on high-density lipoprotein (HDL) cholesterol 4 years post-delivery (**Table S5**). The peak effect at the 11^th^ month after LMP was associated with a 0.07 mg/dL decrease in HDL (95% CI: -0.11, -0.02) per μg/m^3^ increase in PM_2.5_. In addition, we observed associations between increased PM_2.5_ exposure throughout preconception, pregnancy, and the post-partum period (2 months prior to and up to month 17 after LMP) and higher low-density lipoprotein (LDL) cholesterol at the 4-year post-delivery visit. The largest associations occurred during pre-conception (2 months prior to conception) which led to an increase of 0.17 mg/dL in LDL (95% CI: 0.00, 0.34) per PM_2.5_ μg/m^3^ (**Table S6**). While no critical windows of susceptibility to PM_2.5_ were observed with respect to total cholesterol, the overall trend was positive in directionality between the pre-conception and pregnancy PM_2.5_ exposure periods with cholesterol levels at the 4-year post-delivery visit that attenuated over time.

Compared to main results, sensitivity analyses that did not control for pre-pregnancy BMI showed consistent windows of susceptibility to PM_2.5_ exposure across health outcomes and across follow-up visits (**Figures S4-S5**). Also, no notable differences were observed in sensitivity models that controlled for time-varying 1-year PM_2.5_ exposures average prior to follow-up visits (**Figures S6-S7**), or without time-varying pregnancies as covariate (**Figures S8-S9**). Stratified analyses by fetal sex indicated stronger effects positive in directionality on anthropometric markers in pregnant mothers carrying a male fetus compared to mothers carrying a female fetus (**Figures S10-S12**), particularly for WC (**Figure S11**). On the other hand, stratified analyses by fetal sex showed more persistent effects from PM_2.5_ exposures during pregnancy on lipids over time in pregnant mothers carrying a female fetus (**Figures S13-S15**). For instance, windows of susceptibility to PM_2.5_ on higher levels of LDL were sustained throughout all follow-up visits in pregnant mothers carrying a female fetus whereas the associations disappeared over time in those carrying a male fetus (**Figure S15**). Interestingly, stratified analyses by FA intake showed more prominent deleterious effects in the low FA intake group as compared to the high FA intake group (**Figures S16-S21**). This is apparent for all anthropometric outcomes but particularly for WC and body fat where women with lower FA intake levels during pregnancy had lagged PM_2.5_-induced increased levels of adiposity as compared to women with higher FA intake levels (**Figures S17-S18**). Similarly, it is notable that windows of susceptibility to PM_2.5_ on higher LDL and lower HDL cholesterol were stronger in women with lower FA intake levels than in their counterparts (**Figures S20-S21**).

## 4. Discussion

This is the first study assessing long-term alterations of maternal adiposity and lipid measures stemming from PM_2.5_ exposures occurring during and surrounding the time of pregnancy (peri-pregnancy). Several maternal biomarkers assessed at the 4- and 6-year post-delivery visits showed potentially deleterious associations from PM_2.5_ exposures during the period surrounding pregnancy. Long-term adverse associations induced by pregnancy PM_2.5_ exposure were particularly observed for WC (at the 6 year post-delivery visit) and for lipids (at the 4 year post-delivery visit). Critical PM_2.5_ exposure windows were identified during mid and late pregnancy for WC, whereas for lipid outcomes sensitive windows were prominent throughout all pregnancy and during the first post-partum months. Stratified analyses by fetal sex indicated potential stronger deleterious effects with anthropometric measures that were more prominent in mothers carrying a male fetus, whereas PM_2.5_ associations with lipid biomarkers were stronger in mothers carrying a female fetus. Stratified analyses indicated also potential stronger deleterious effects in women with lower FA intake during pregnancy.

In this study, we showed that PM_2.5_ exposures during and surrounding pregnancy could exacerbate potential physiological changes occurring during gestation and we also showed that those associations can last up to 6 years post-delivery. Pregnancy induces physiological changes that uniquely affect maternal metabolism influencing post-partum weight retention.^83,84^ Weight gain can be sustained after delivery through weight retention triggered by pro-inflammatory processes starting during conception.^85–87^ For instance, during gestation major changes in lipid and glucose homeostasis ensure an adequate supply of energy to the developing fetus.^88,89^ These changes could promote body fat accumulation,^90,91^ changes in leptin hormone levels,^92^ increased glucose production,^93^ which in turn, can lead to increased risk of developing gestational diabetes^94^ and subsequent type-2 diabetes or cardiovascular diseases later in life.^49,50^ Increased adiposity has been previously observed in adults exposed to PM_2.5_.^95-97^

PM_2.5_ exposures during mid-pregnancy altered cholesterol levels up to the 4-year post-delivery visit. Our findings showed that pregnancy and early post-partum PM_2.5_ exposure increased levels of LDL and decreased levels of HDL. These PM_2.5_-associated increases in LDL levels or decreases in HDL levels in women having undergone pregnancy were consistent with previous literature in non-pregnant adults.^28,98^ Higher LDL and lower HDL cholesterol levels have been shown to predispose women to higher risk of cardiovascular events later in life.^99–101,102,103^ Currently, there is a scarcity of studies examining lipid levels in pregnant women that also examine critical periods of susceptibility to PM_2.5_ but one lipidomic study indicated that PM_2.5_ during the 2^nd^ trimester can exacerbate adverse birth outcomes in women with hyperlipidemia through increased inflammation.^104^

Our findings also suggested that pregnancy PM_2.5_ exposures are associated with adiposity and adverse lipid alterations up to 6 years post-delivery but not at 8 years in our sample. It is possible that PM_2.5_ exposures during and surrounding pregnancy may still have lingering deleterious effects at later stages in life but subsequent exposures could also be inflicting adverse effects and drown out the signal of more distant exposure such as those occurring in pregnancy years prior. Moreover, future studies with longer follow-up time are needed to elucidate whether protracted effects from pregnancy PM_2.5_ exposure go beyond 6 years post-delivery. To evaluate the potential influence of concurrent PM_2.5_ exposures, we adjusted for average PM_2.5_ exposures throughout a 1-year span prior to each examination visit as a time-varying variable in sensitivity analyses, which did not change the results of primary analyses. Although differences in associations over time may reflect women’s lifestyle changes and/or weight changes after giving birth, we controlled for several time-varying lifestyle variables in analyses. In addition, we did not observe clear patterns of associations between lagged PM_2.5_ exposures throughout pregnancy and anthropometric markers other than WC, suggesting that protracted PM_2.5_ exposures may have later effects on abdominal adiposity but perhaps not on generalized adiposity in birthing people, which is in line with the absence of associations with total body fat in our study.

We also observed potential diverging effects by fetal sex in our study. Critical windows of susceptibility to pregnancy PM_2.5_ exposure may be sex-dimorphic due to different circulating levels of sex-steroid hormones during pregnancy^79,80^ that may exacerbate or alter effects from air pollution. Our findings are consistent with previous literature indicating that carrying a male fetus is accompanied with a higher risk of poor cardiovascular health for the mother resulting in pregnancy complications and/or adverse health.^54^ Moreover, our results indicated also a suggestive protective role of FA against protracted effects from PM_2.5_ exposure. These results are consistent with prior findings in PROGRESS mothers showing beneficial effects of higher FA intake levels (above 600 mcg/day) during pregnancy in the associations of air pollutant mixtures and risk of liver injury.^81^ In addition to cardiometabolic outcomes, FA has been shown in prior literature to attenuate the risk of adverse birth outcomes induced by airborne particulate matter during pregnancy.^55^ However, these hypotheses suggested in stratified analyses warrant further exploration and need to be formally tested with new methods accommodating for distributed lag models that can handle multiple modifying factors.

We also recognize potential limitations in our study. First, estimates for months 7 through 9 after LMP refer to a population at the 3^rd^ trimester of gestation but also encompasses a small fraction of women (n=55; ∼11%) with early-term pregnancies (baby born before the 37^th^ week of pregnancy). Second, our study intended to focus specifically in the most relevant epidemics of generalized and abdominal adiposity in the Mexican population, as well as its top major dyslipidemias: hypoalphalipoproteinemia and hypercholesterolemia. However, other markers of metabolic syndrome such as blood pressure and diabetes-markers may be of interest to explore in future studies. We also acknowledge potential measurement error from self-reported covariates and underreport of compromising lifestyle information during pregnancy such as alcohol intake and smoking.^105,106^ While we acknowledge that measurement errors of the exposure modeling can introduce some degree of exposure misclassification, we believe that our overall approach— including the use of validated models, cross-validation, and conservative monthly exposure averaging— reduces these errors and enhances the reliability of the estimates. We also used the first day of the LMP as a proxy for conception, which can be prone to biological error.^107,108^ However, we do not expect a substantial change in the interpretation as it was conducted in monthly intervals, thus minimizing the biological error, and we used a validated approach to identify LMP when gestational age differed by more than three weeks to LMP (**Methods S2**).^62^ In addition, due to the poor analytical performance of LDL-C analysis methods,^109,110^ no direct LDL measurement in blood was performed in our study. Although a comprehensive adjustment of covariates was implemented, unmeasured confounding biasing results could also still be present as this is an observational study. Given that it is an observational study, we recognize that we cannot directly establish a causal relationship between critical windows to PM_2.5_ exposure on maternal lipids and waist circumference and that instead we are examining associations. Similarly, we did not measure other concurrent environmental exposures that could be correlated with PM_2.5_ driving the associations. Also, our effect estimates are small and results should be interpreted with caution. Last, although this study in Mexican women may not be generalizable to other populations, using a relatively homogenous sample increases the internal validity of our results.

Our study also benefits from several strengths. To our knowledge, this is the first study assessing the long-term cardiometabolic effects of pregnancy PM_2.5_ exposure in a Hispanic population and using a comprehensive panel of anthropometric and lipids. Our study also uses state-of-the-art methods to characterize daily PM_2.5_ air pollution data. We also used a novel application of a recently developed statistical model to simultaneously estimate critical windows of exposure in a trajectory analysis with repeated outcome assessment and a continuous time component. This has the advantage to reduce bias using the DLM framework^111^ while pooling strength and gaining power from repeated outcome assessments. We also leveraged a population of pregnant women from Central Mexico exposed to high PM_2.5_ levels that could be more susceptible to developing cardiometabolic diseases. In Mexico, the prevalence of overweight and obesity reached 39% in 2018 with mid-life adults, and women in particular, having the highest rates.^112^ Our study also highlights that in Mexico City average PM_2.5_ levels are 4-5 times the WHO recommended annual average levels (5 μg/m^3^),^113^ making these findings of interest to help drive policy change, and particularly in view of the UN Sustainable Development Goals aiming to prevent non-communicable diseases in lower-middle-income countries.^114^ Additional studies should be conducted in pregnant women with more than eight years of follow-up, using multi-pollutant protracted exposures, directly measuring LDL via centrifugation or homogenous assays,^109,115^ leveraging different populations outside Mexico, and evaluating high-order interactions with effect modifiers. Our study provides a framework for assessing windows of susceptibility with trajectory outcomes that can be replicated in such future studies.

## 5. Conclusion

Pregnancy itself may constitute a critical window to PM_2.5_ exposure for dysregulated long-term maternal metabolic health after delivery. Pregnant women could largely benefit from policies targeting reduction to air pollution exposure.

## 6. Data Sharing

Data is only available upon reasonable request from the authors.

## 8. Acknowledgements

The work involved in this study was also supported by UL1TR004419 (PI: Wright), P30ES023515 (PI: Wright), R01ES029943 (PI: Kioumourtzoglou, Weisskopf), R01ES033688 (PI: Valvi), R21ES035148 (PI: Valvi).

## 9. Author Contributions

E.C., A.W.: Conceptualization; S.I.A., D.D.: Data curation, Formal analysis, Investigation; E.C., H.W., A.A.B.: Funding acquisition; S.I.A., D.D.: Software, Visualization; S.I.A., D.D., E.C., A.W., K.K.: Methodology; S.I.A.: Roles/Writing–original draft; D.D., D.V., A.C.J., I.G.A., G.E., M.M.T.R., R.O.W., A.A.B., H.W., K.P.K., A.W., E.C.: Writing—review & editing.

