## Supplementary Material 1 for "Windows of Susceptibility to Air Pollution During and Surrounding Pregnancy in Relation to Longitudinal Maternal Measures of Adiposity and Lipid Profiles"

**Supplemental Methods 1.**

Our model was evaluated with leave-one-station-out cross-validation (CV), and exhibited good performance, with an overall cross-validated mean absolute error of 3.68 μg/m^3^ and an annual R^2^ ranging from 0.64 to 0.86. We estimated residential exposure to PM_2.5_ at each participant's address, accounting for any moves, as follows. Participants’ addresses were collected at recruitment and updated at each follow-up if they had moved, as well as through an address history questionnaire administered between 2018-2020. Residential addresses were geocoded and crosschecked against actual geographic coordinates collected by our fieldwork team at each study visit. PM_2.5_ exposure estimates were assigned to study participants based on the geographic coordinates of their homes, using the corresponding 1 × 1 km PM_2.5_ grid cells from our satellite-based models.^1^

**Supplemental Methods 2.**

We flagged self-reported medications to treat cardiometabolic disease^2^ at baseline and at each follow-up visit. These included self-report of a drug treatment for abnormal levels of either glucose, triglycerides, high-density lipoprotein (HDL), or blood pressure. Given that participants self-reported their specific medications at the visit by filling out a questionnaire with blank spaces with the name of the medication, only a list of self-reported names of medications are provided by the participants and limited information exists on whether healthy individuals without reported medications are non-respondents. The list of medications self-reported by participants across all visits were the following (in alphabetical order):

- Acarbose
- Alpha-methyldopa
- Amlodipine
- Atorvastatin
- Bezafibrate
- Captopril
- Chlortalidone
- Enalapril
- Farxiga
- Glibenclamide
- Hydrochlorothiazide
- Insulin
- Januvia
- Kombiglyze
- Losartan
- Pravastatin
- Propranolol
- Methyldopa
- Metformin
- Metoprolol
- Norvasc
- Telmisartan
- Verapamil

**Supplemental Methods 3.**

The distributed lag interaction model (DLIM) estimates a linear exposure-time-response function that varies continuously by time after delivery and by outcome assessment time. The DLIM is

$$y_{ij}=\sum_{t=-2}^{22} x_{it}\beta_{t}(m_{ij})+\boldsymbol{z}_{ij}^{'}\boldsymbol{\gamma}+b_{i}+\varepsilon_{ij},$$

where $y_{ij}$is the $j$th outcome for individual $i$, $x_{it}$ is the exposure level for individual $i$ in month $t$, $\beta_{t}(m_{i})$ is the coefficient for the linear effect of exposure at time $t$ for an individual with outcome assessed at time $m_{ij}$. The outcome time $m_{ij}$ is in months since delivery rounded to the first decimal place of the $j$th post-delivery visit and, therefore, accounts for deviations in visit time from the 4, 6, and 8-year targets and allow for estimation of the lagged effects at any time during the observed outcome time span. In addition, the term $z_{ij}^{'}\gamma$ controls for covariates, $b_{i}$ is an individual random intercept to account for correlation between repeated measures for an individual, and $\varepsilon_{ij}$ is a residual error term. To construct the cross-basis for the DLIM, we used penalized B-splines^3^ with 20 basis functions to smooth in both the exposure-time and post-delivery-time dimensions. We estimated the model parameters with restricted maximum likelihood as previously described.^4^ We performed bootstrap likelihood ratio tests to choose the basis construction, linear modification, non-linear modification, or no modification.^4^

**Supplemental Figure 1. Study Participation Flowchart.**

n=1054 mothers

n=948 with live births or no loss to follow-up during pregnancy

n=810 with air pollution data

n=806 with no missing information on pre-pregnancy BMI, age, and seasonality

n=517 with at least 1 anthropometry or cholesterol measure between follow-up visit at 48 months and 96 months

**Supplemental Figure 2. Directed Acyclic Graph (DAG) between PM_2.5_ and Cardiometabolic Health. ^a^**

Age

Pre-pregnancy BMI

Socio-economic Status (SES)

Marital Status

Parity at Enrollment

(-)

^a^ Includes potential confounders (seasonality, SES) and additional clinically relevant predictors (alcohol, smoking, cardiometabolic medications, parity, age, pre-pregnancy BMI, marital status, additional pregnancies during follow-up, and stage).

(-)

(-)

(+)

(+)

(-)

(+)

(-)

Season (Dry/Warm)

Cardiometabolic Medications

Alcohol Intake

Smoking during Pregnancy

Multiple Pregnancies

Follow-Up Visit (Stage)

(+)

(-)

(-)

(+)

(+)

PM_2.5_

Cardiometabolic Health

**Supplemental Figure 3. PM_2.5_ Exposure Levels during and around Pregnancy from 2 Months Prior to Last Menstrual Period (LMP) to 22 Months after LMP.**

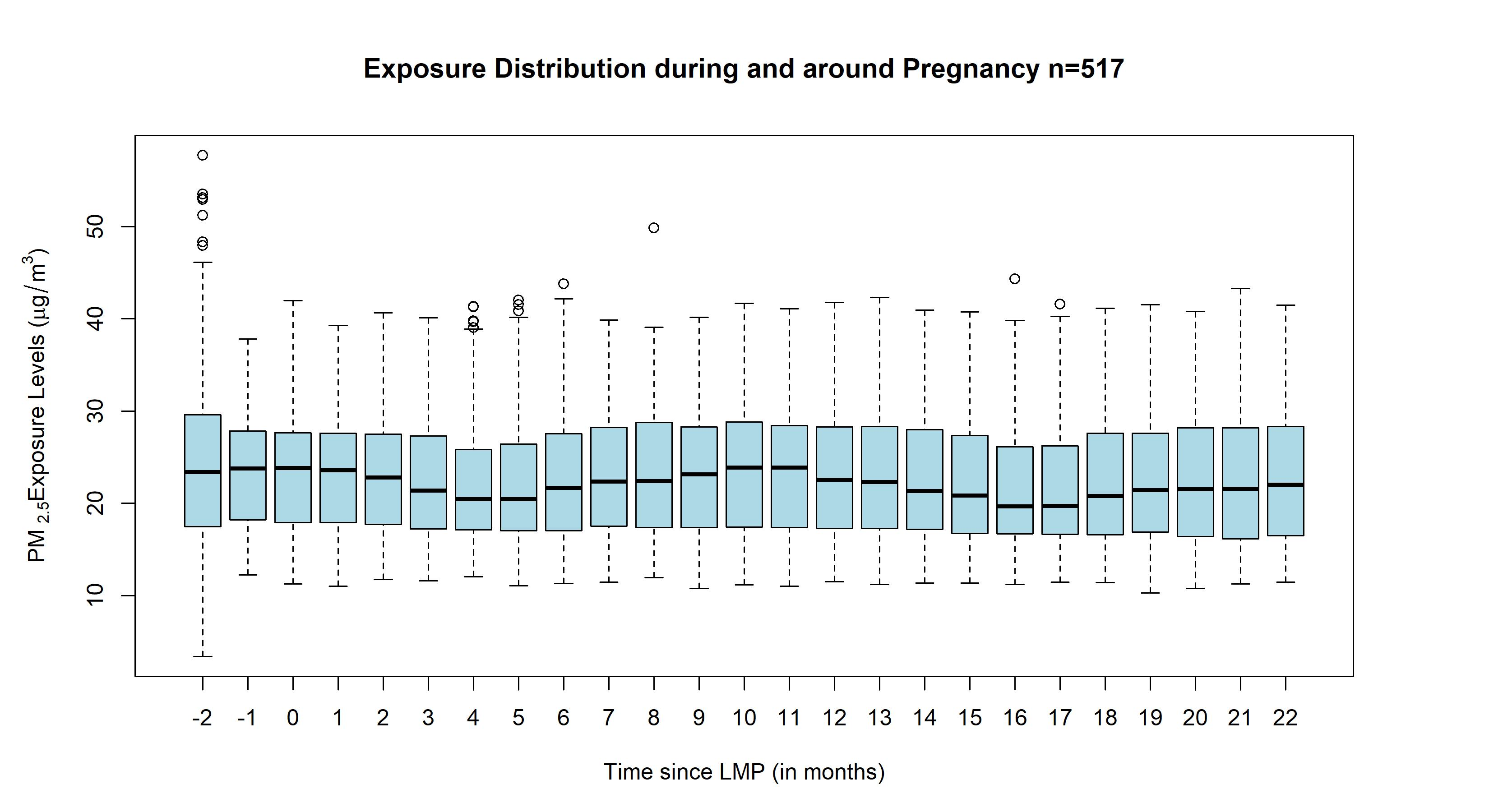

**Supplemental Figure 4.** Sensitivity analyses for anthropometric measures showing overlaying plots with original distributed lag interaction models (DLIMs) controlling for pre-pregnancy body mass index (BMI) in gray color and DLIMs not controlling for BMI as a covariate in blue color. All models were additionally adjusted for age, socio-economic status, smoking during pregnancy, marital status, parity at enrollment, meteorological season, cardiometabolic medications, alcohol intake, multiple pregnancies throughout follow-up, stage, and follow-up time in months squared to allow for a nonlinear effect. Estimates of the association and 95% confidence interval (95% CI) between monthly PM_2.5_ exposure and anthropometric measures Body Mass Index (BMI), Waist Circumference (WC), body fat percentage at 48, 72, and 96 months after parturition using DLIMs with penalization and non-linear modification. Negative and positive values on the x-axis indicate months before and after last menstrual period (LMP), respectively. Month 0 indicates the LMP month (month 0). Sample sizes for analyses with BMI were n=516 and sample sizes for analyses with WC and body fat were n=515.

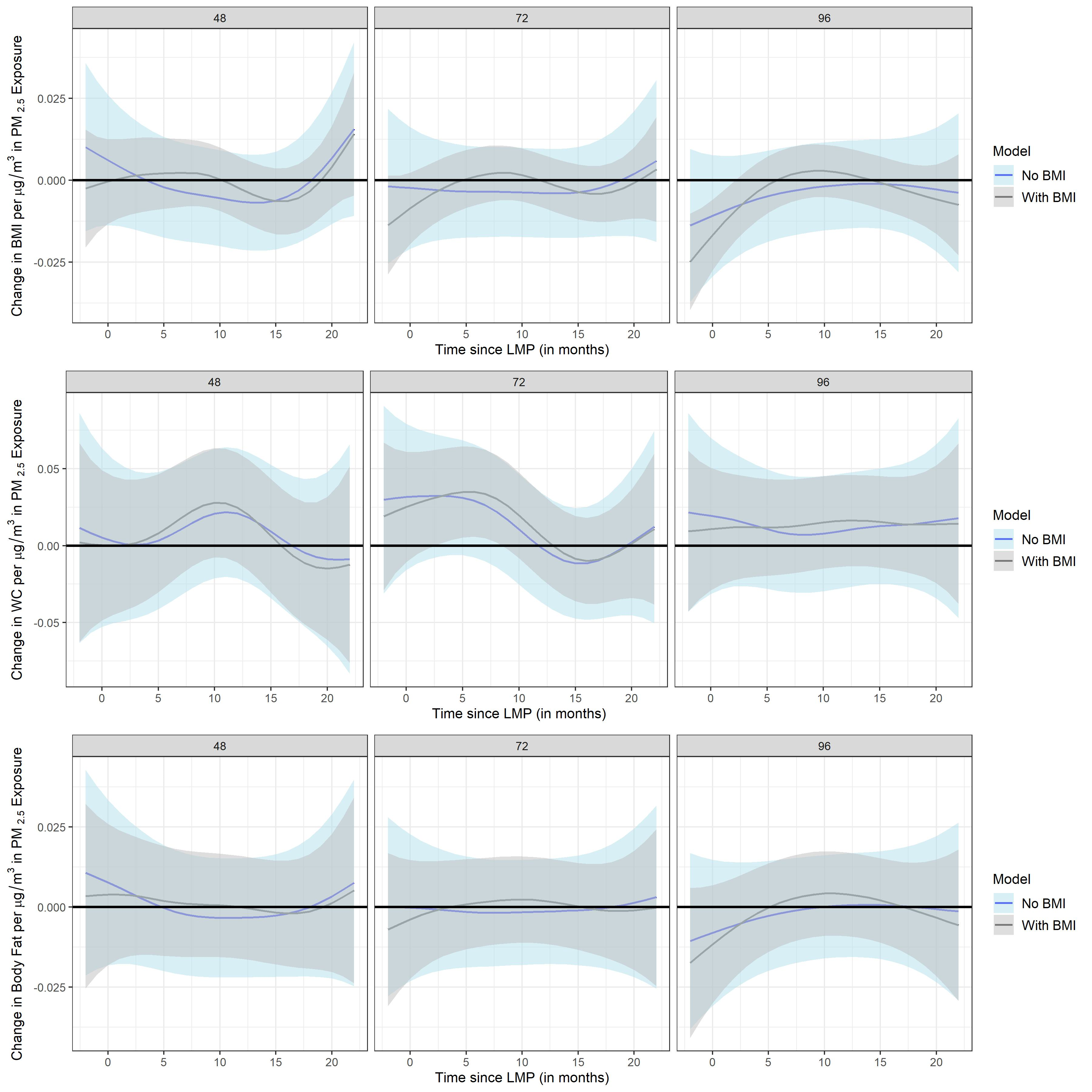

**Supplemental Figure 5.** Sensitivity analyses for cholesterol biomarkers showing overlaying plots with original distributed lag interaction models (DLIMs) controlling for pre-pregnancy body mass index (BMI) in gray color and DLIMs not controlling for BMI as a covariate in blue color. All models were additionally adjusted for age, socio-economic status, smoking during pregnancy, marital status, parity at enrollment, meteorological season, cardiometabolic medications, alcohol intake, multiple pregnancies throughout follow-up, stage, and follow-up time in months squared to allow for a nonlinear effect. Estimates of the association and 95% confidence interval (95% CI) between monthly PM_2.5_ exposure and total cholesterol, high-density lipoprotein (HDL), low-density lipoprotein (LDL) at 48, 72, and 96 months after parturition using DLIMs with penalization and non-linear modification. Negative and positive values on the x-axis indicate months before and after last menstrual period (LMP), respectively. Month 0 indicates the LMP month (month 0). Sample sizes for analyses with total cholesterol, HDL, and LDL were n=517.

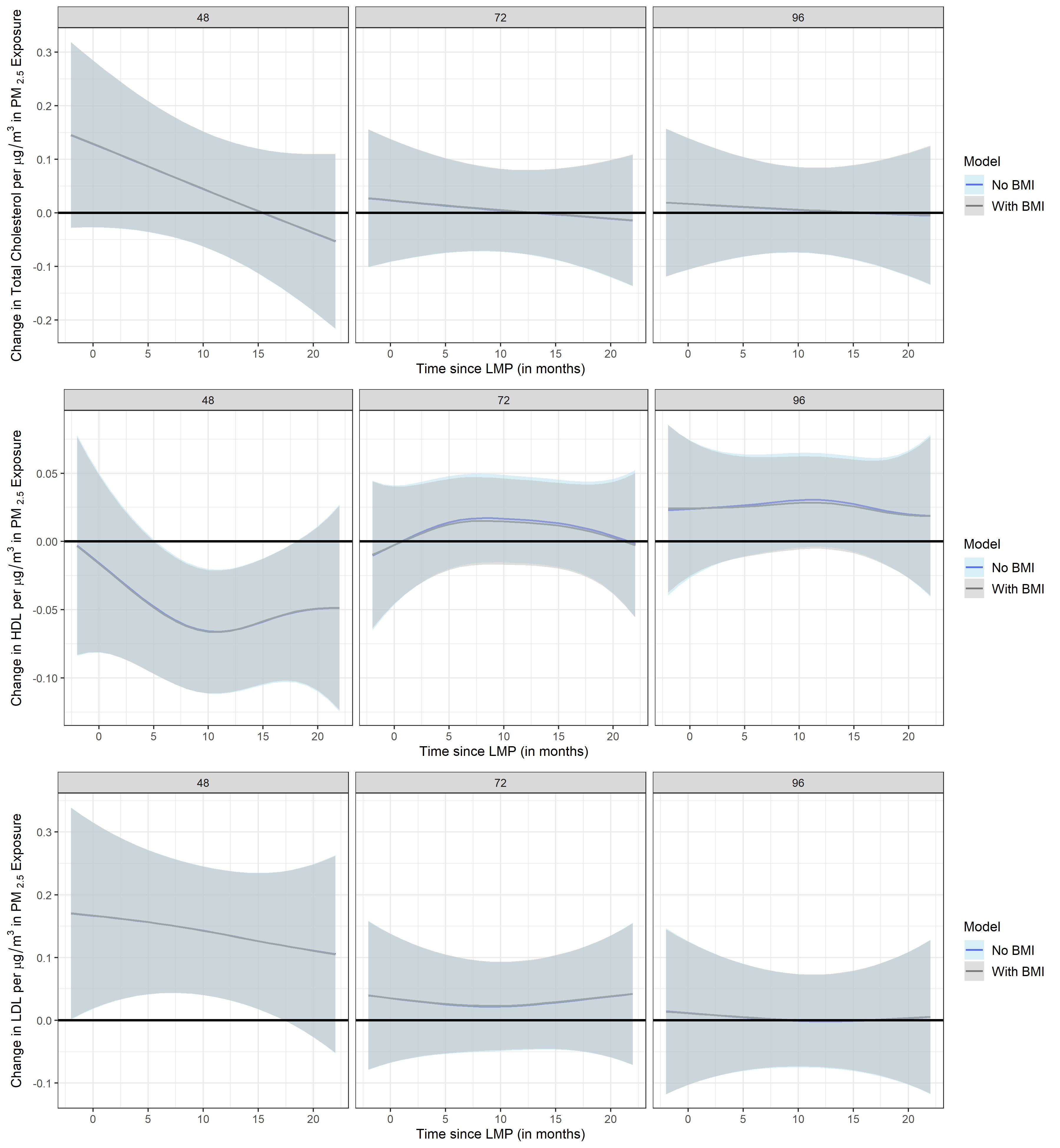

**Supplemental Figure 6.** Sensitivity analyses for anthropometric measures showing overlaying plots with original distributed lag interaction models (DLIMs) not controlling for time-varying 1-year average PM_2.5_ (prior to each follow-up visit) in gray color and DLIMs controlling for time-varying 1-year average PM_2.5_ as a covariate in blue color. All models were additionally adjusted for age, pre-pregnancy BMI, socio-economic status, smoking during pregnancy, marital status, parity at enrollment, meteorological season, cardiometabolic medications, alcohol intake, multiple pregnancies throughout follow-up, stage, and follow-up time in months squared to allow for a nonlinear effect. Estimates of the association and 95% confidence interval (95% CI) between monthly PM_2.5_ exposure and anthropometric measures Body Mass Index (BMI), Waist Circumference (WC), body fat percentage at 48, 72, and 96 months after parturition using DLIMs with penalization and non-linear modification. Negative and positive values on the x-axis indicate months before and after last menstrual period (LMP), respectively. Month 0 indicates the LMP month (month 0). Sample sizes for analyses with BMI were n=516 and sample sizes for analyses with WC and body fat were n=515.

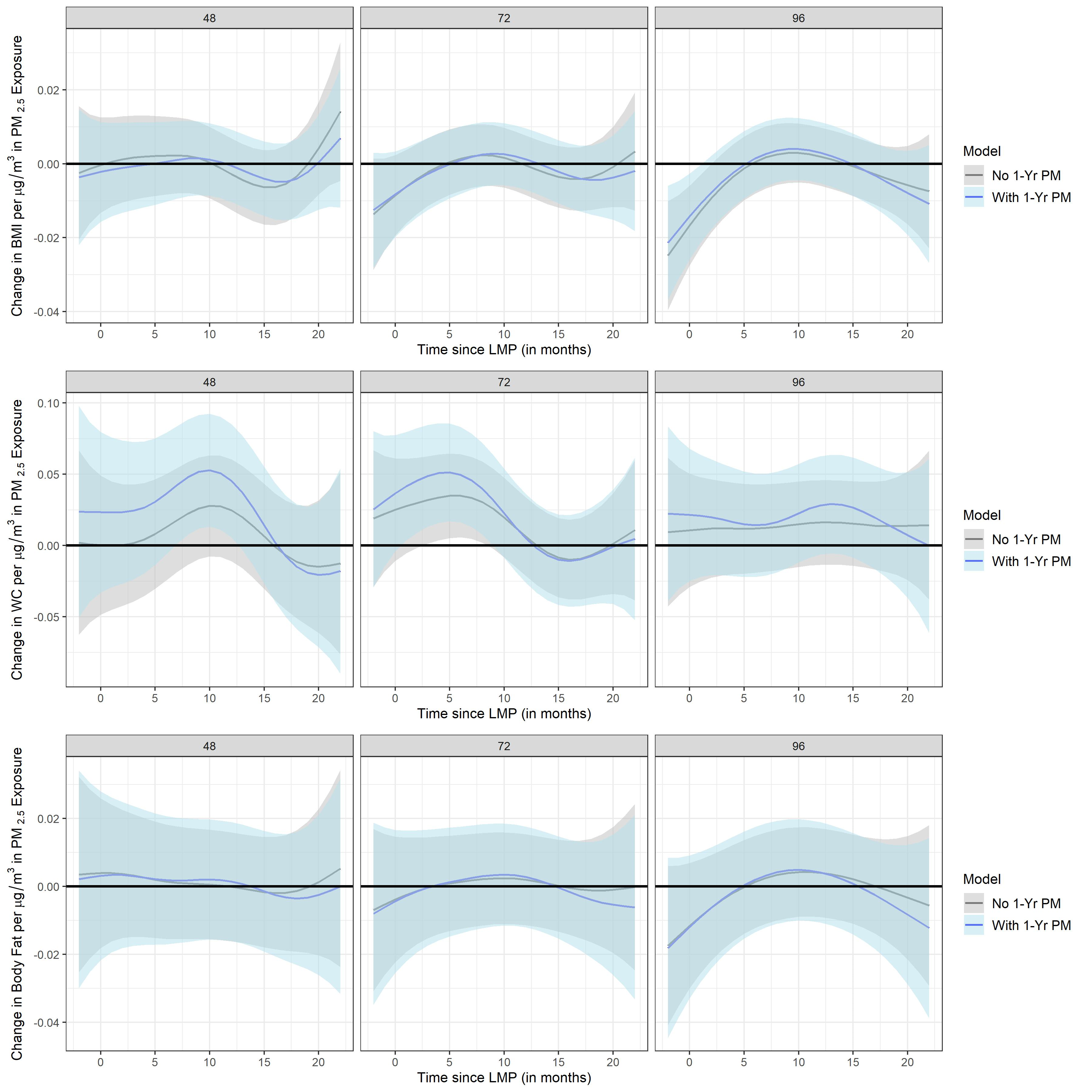

**Supplemental Figure 7.** Sensitivity analyses for cholesterol biomarkers showing overlaying plots with original distributed lag interaction models (DLIMs) not controlling for time-varying 1-year average PM_2.5_ (prior to each follow-up visit) in gray color and DLIMs controlling for time-varying 1-year average PM_2.5_ as a covariate in blue color. All models were additionally adjusted for age, pre-pregnancy BMI, socio-economic status, smoking during pregnancy, marital status, parity at enrollment, meteorological season, cardiometabolic medications, alcohol intake, multiple pregnancies throughout follow-up, stage, and follow-up time in months squared to allow for a nonlinear effect. Estimates of the association and 95% confidence interval (95% CI) between monthly PM_2.5_ exposure and total cholesterol, high-density lipoprotein (HDL), low-density lipoprotein (LDL) at 48, 72, and 96 months after parturition using DLIMs with penalization and non-linear modification. Negative and positive values on the x-axis indicate months before and after last menstrual period (LMP), respectively. Month 0 indicates the LMP month (month 0). Sample sizes for analyses with total cholesterol, HDL, and LDL were n=517.

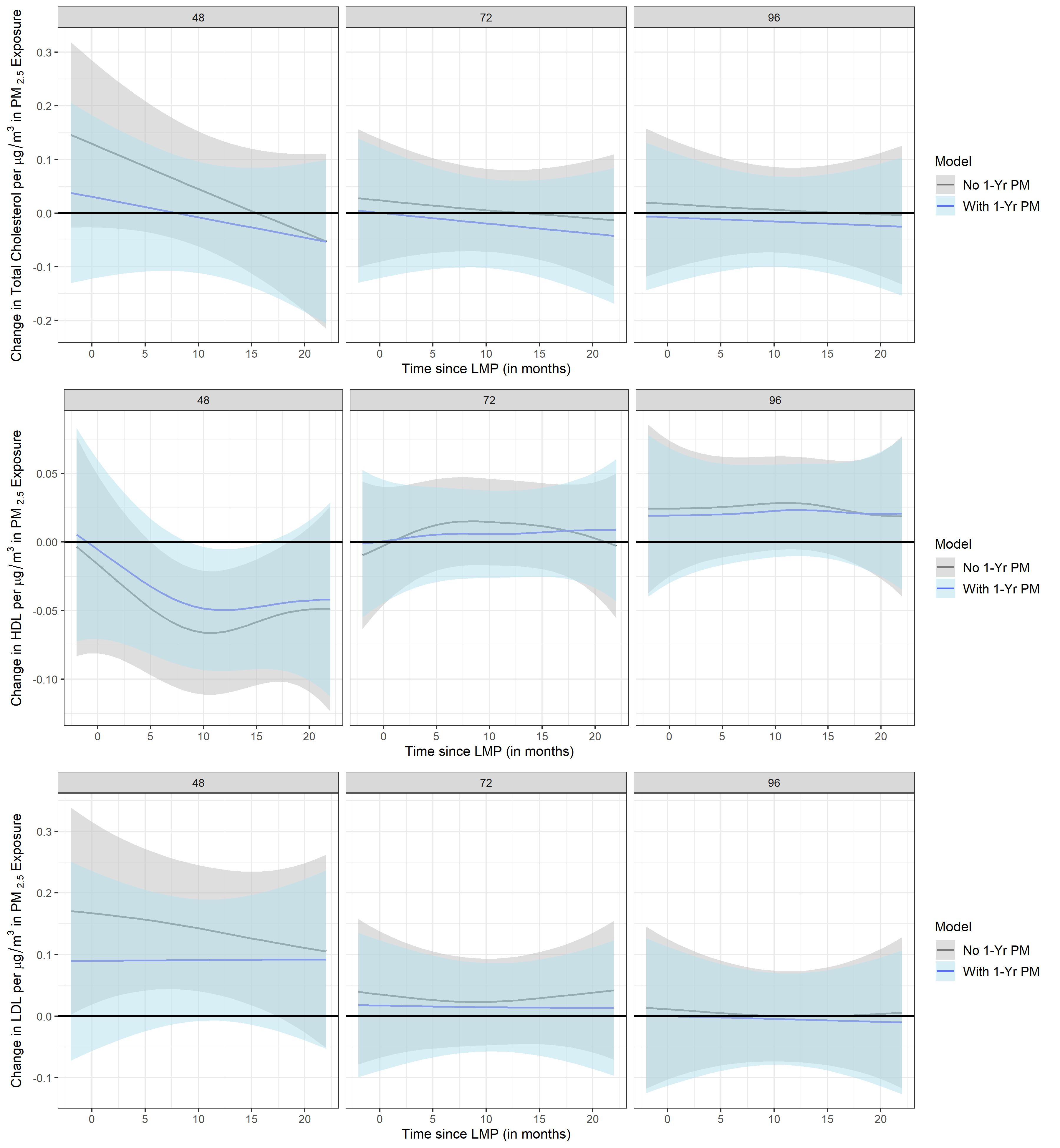

**Supplemental Figure 8.** Sensitivity analyses for anthropometric measures showing overlaying plots with original distributed lag interaction models (DLIMs) controlling for time-varying pregnancies as a covariate in gray color and DLIMs not controlling for time-varying pregnancies in blue color. All models were additionally adjusted for age, pre-pregnancy BMI, socio-economic status, smoking during pregnancy, marital status, parity at enrollment, meteorological season, cardiometabolic medications, alcohol intake, stage, and follow-up time in months squared to allow for a nonlinear effect. Estimates of the association and 95% confidence interval (95% CI) between monthly PM_2.5_ exposure and anthropometric measures Body Mass Index (BMI), Waist Circumference (WC), body fat percentage at 48, 72, and 96 months after parturition using DLIMs with penalization and non-linear modification. Negative and positive values on the x-axis indicate months before and after last menstrual period (LMP), respectively. Month 0 indicates the LMP month (month 0). Sample sizes for analyses with BMI were n=516 and sample sizes for analyses with WC and body fat were n=515.

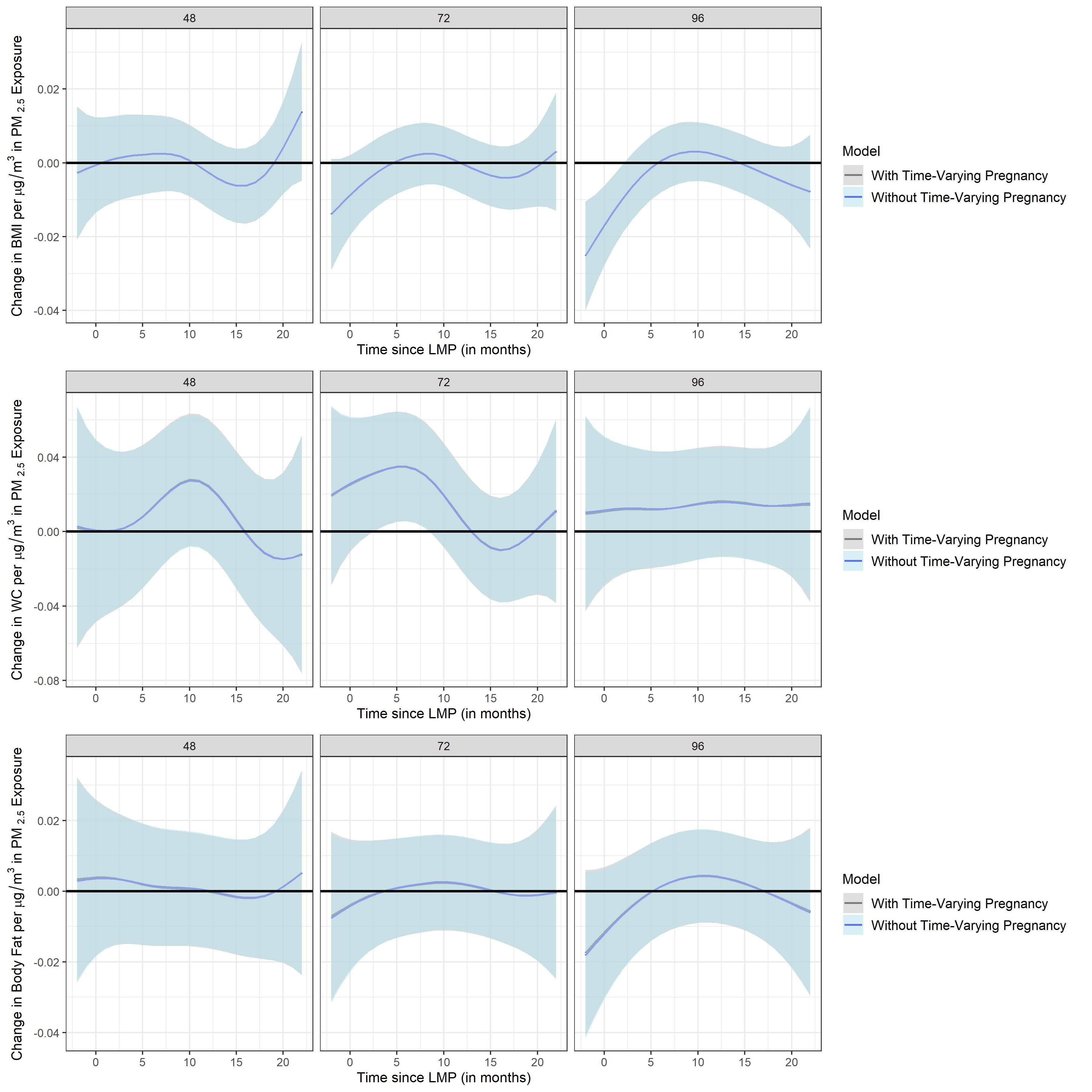

**Supplemental Figure 9.** Sensitivity analyses for cholesterol biomarkers showing overlaying plots with original distributed lag interaction models (DLIMs) controlling for time-varying pregnancies as a covariate in gray color and DLIMs not controlling for time-varying pregnancies in blue color. All models were additionally adjusted for age, pre-pregnancy BMI, socio-economic status, smoking during pregnancy, marital status, parity at enrollment, meteorological season, cardiometabolic medications, alcohol intake, stage, and follow-up time in months squared to allow for a nonlinear effect. Estimates of the association and 95% confidence interval (95% CI) between monthly PM_2.5_ exposure and total cholesterol, high-density lipoprotein (HDL), low-density lipoprotein (LDL) at 48, 72, and 96 months after parturition using DLIMs with penalization and non-linear modification. Negative and positive values on the x-axis indicate months before and after last menstrual period (LMP), respectively. Month 0 indicates the LMP month (month 0). Sample sizes for analyses with total cholesterol, HDL, and LDL were n=517.

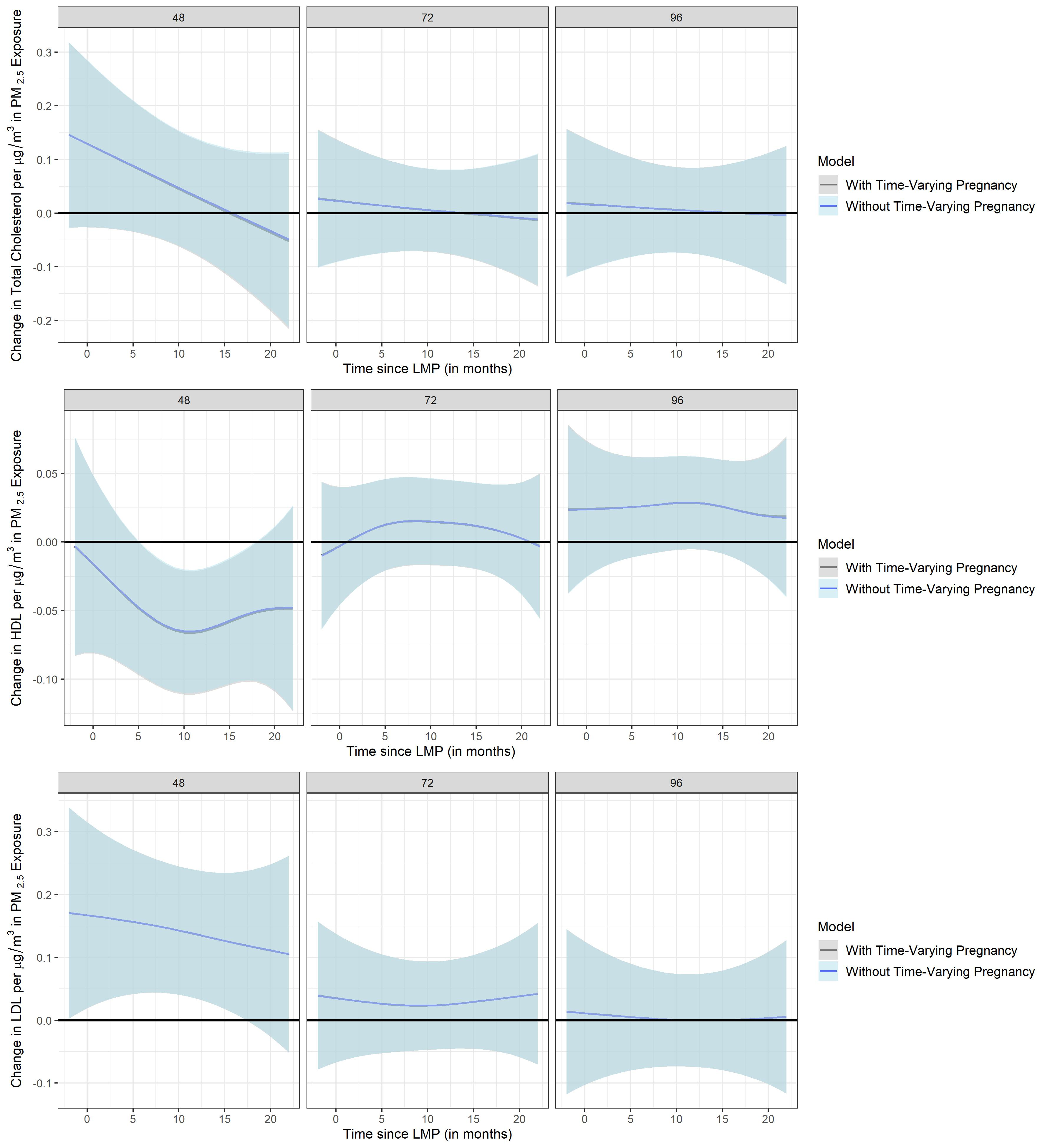

**Supplemental Figure 10.** Stratified analyses by fetal sex (pregnant mothers carrying either a male or female fetus) using distributed lag interaction models (DLIMs) with penalization and non-linear modification. Estimates of the association (blue dots) and 95% confidence interval (95% CI) (grey whiskers) between monthly PM_2.5_ exposure and Body Mass Index (BMI) at 48, 72, and 96 months after parturition. Negative and positive values on the x-axis indicate months before and after last menstrual period (LMP), respectively. Month 0 indicates the LMP month (month 0). Models were adjusted for age, pre-pregnancy BMI, socio-economic status, smoking during pregnancy, marital status, parity at enrollment, meteorological season, cardiometabolic medications, alcohol intake, multiple pregnancies throughout follow-up, stage, and follow-up time in months squared to allow for a nonlinear effect. Sample sizes for stratified analyses in pregnant mothers were n=267 for the male fetus carrier group and n=249 for the female fetus carrier group.

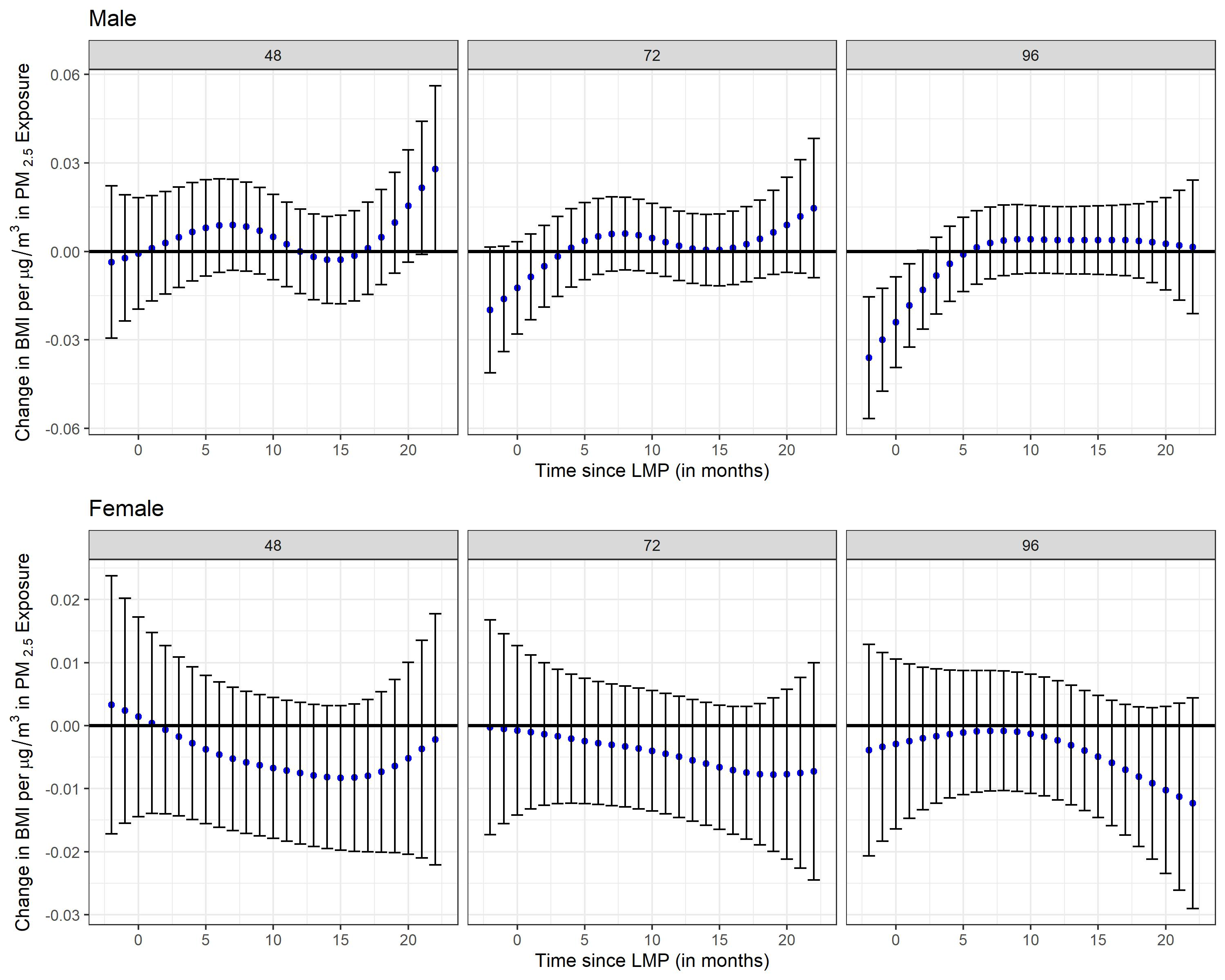

**Supplemental Figure 11.** Stratified analyses by fetal sex (pregnant mothers carrying either a male or female fetus) using distributed lag interaction models (DLIMs) with penalization and non-linear modification. Estimates of the association (blue dots) and 95% confidence interval (95% CI) (grey whiskers) between monthly PM_2.5_ exposure and Waist Circumference (WC) at 48, 72, and 96 months after parturition. Negative and positive values on the x-axis indicate months before and after last menstrual period (LMP), respectively. Month 0 indicates the LMP month (month 0). Models were adjusted for age, pre-pregnancy BMI, socio-economic status, smoking during pregnancy, marital status, parity at enrollment, meteorological season, cardiometabolic medications, alcohol intake, multiple pregnancies throughout follow-up, stage, and follow-up time in months squared to allow for a nonlinear effect. Sample sizes for stratified analyses in pregnant mothers were n=266 for the male fetus carrier group and n=249 for the female fetus carrier group.

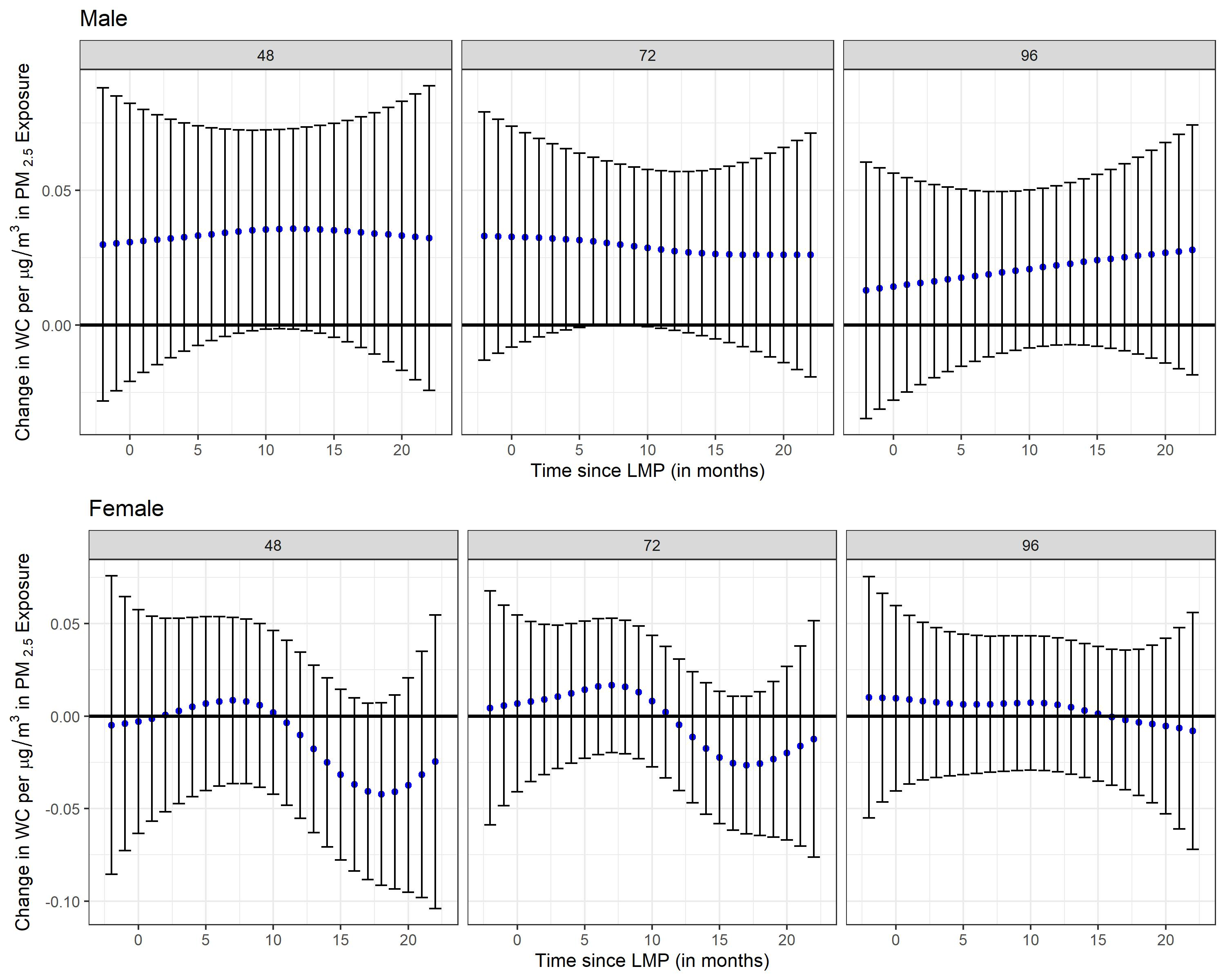

**Supplemental Figure 12.** Stratified analyses by fetal sex (pregnant mothers carrying either a male or female fetus) using distributed lag interaction models (DLIMs) with penalization and non-linear modification. Estimates of the association (blue dots) and 95% confidence interval (95% CI) (grey whiskers) between monthly PM_2.5_ exposure and body fat percentage at 48, 72, and 96 months after parturition. Negative and positive values on the x-axis indicate months before and after last menstrual period (LMP), respectively. Month 0 indicates the LMP month (month 0). Models were adjusted for age, pre-pregnancy BMI, socio-economic status, smoking during pregnancy, marital status, parity at enrollment, meteorological season, cardiometabolic medications, alcohol intake, multiple pregnancies throughout follow-up, stage, and follow-up time in months squared to allow for a nonlinear effect. Sample sizes for stratified analyses in pregnant mothers were n=266 for the male fetus carrier group and n=249 for the female fetus carrier group.

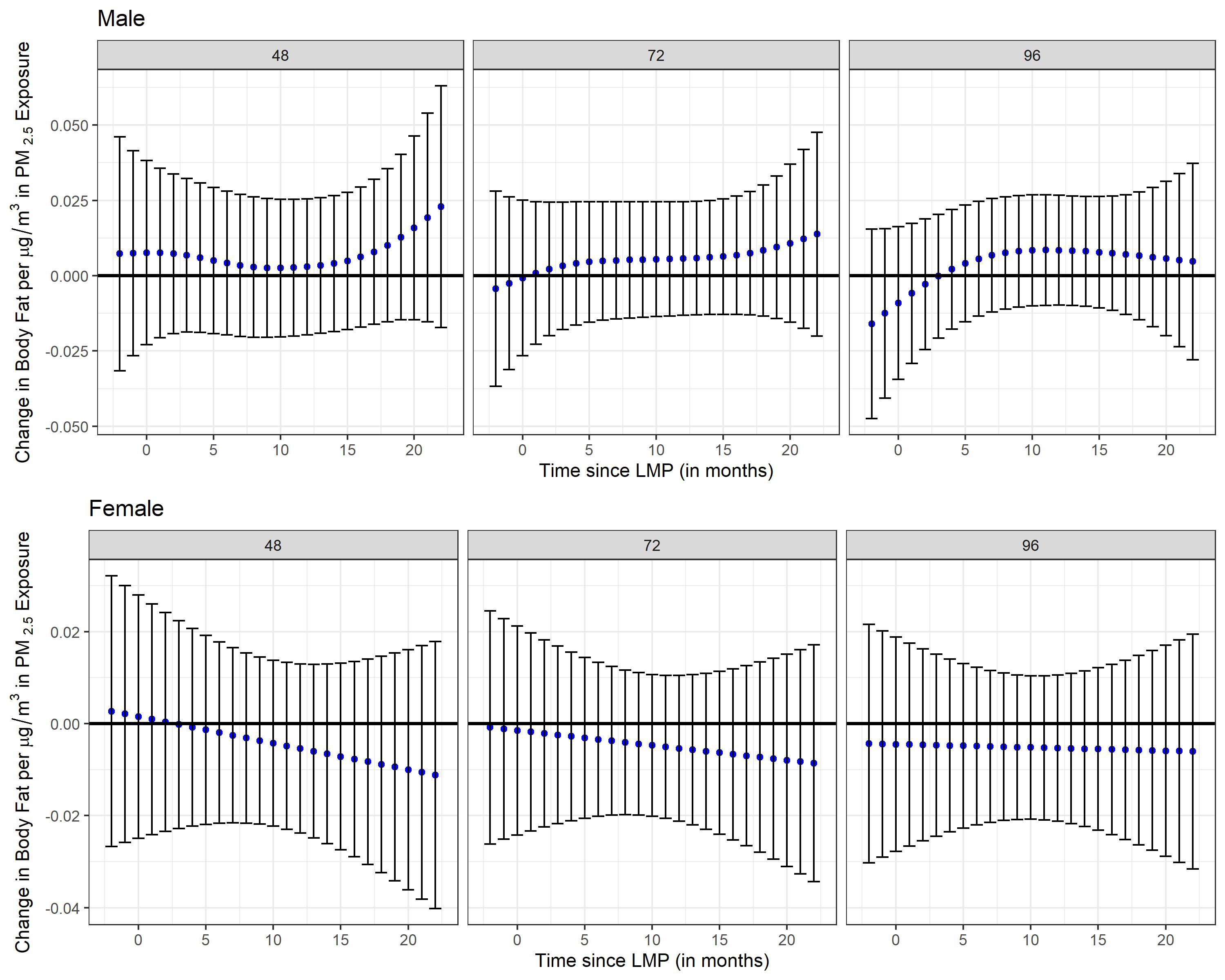

**Supplemental Figure 13.** Stratified analyses by fetal sex (pregnant mothers carrying either a male or female fetus) using distributed lag interaction models (DLIMs) with penalization and non-linear modification. Estimates of the association (blue dots) and 95% confidence interval (95% CI) (grey whiskers) between monthly PM_2.5_ exposure and total cholesterol at 48, 72, and 96 months after parturition. Negative and positive values on the x-axis indicate months before and after last menstrual period (LMP), respectively. Month 0 indicates the LMP month (month 0). Models were adjusted for age, pre-pregnancy BMI, socio-economic status, smoking during pregnancy, marital status, parity at enrollment, meteorological season, cardiometabolic medications, alcohol intake, multiple pregnancies throughout follow-up, stage, and follow-up time in months squared to allow for a nonlinear effect. Sample sizes for stratified analyses in pregnant mothers were n=267 for the male fetus carrier group and n=250 for the female fetus carrier group.

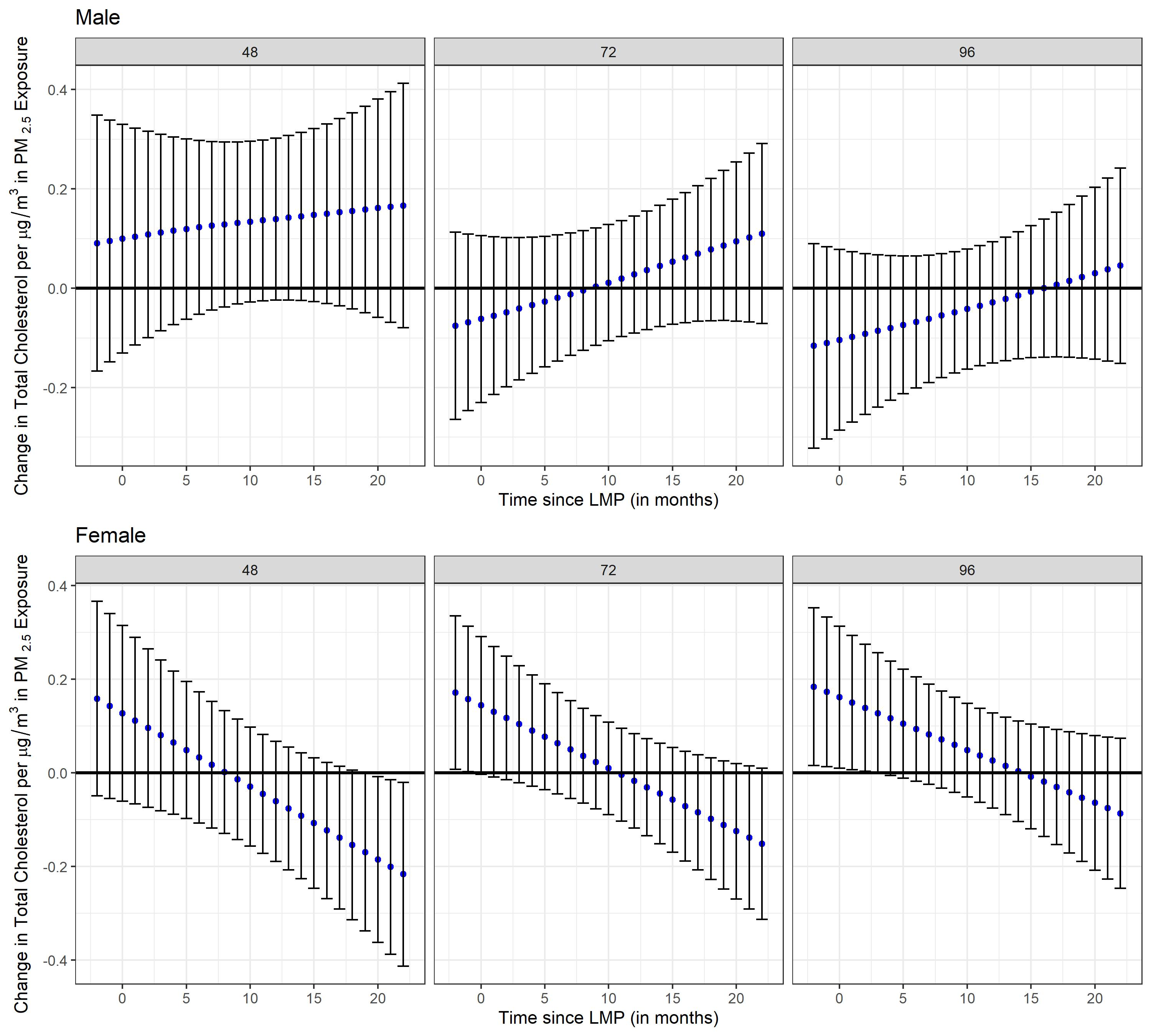

**Supplemental Figure 14.** Stratified analyses by fetal sex (pregnant mothers carrying either a male or female fetus) using distributed lag interaction models (DLIMs) with penalization and non-linear modification. Estimates of the association (blue dots) and 95% confidence interval (95% CI) (grey whiskers) between monthly PM_2.5_ exposure and high-density lipoprotein (HDL) at 48, 72, and 96 months after parturition. Negative and positive values on the x-axis indicate months before and after last menstrual period (LMP), respectively. Month 0 indicates the LMP month (month 0). Models were adjusted for age, pre-pregnancy BMI, socio-economic status, smoking during pregnancy, marital status, parity at enrollment, meteorological season, cardiometabolic medications, alcohol intake, multiple pregnancies throughout follow-up, stage, and follow-up time in months squared to allow for a nonlinear effect. Sample sizes for stratified analyses in pregnant mothers were n=267 for the male fetus carrier group and n=250 for the female fetus carrier group.

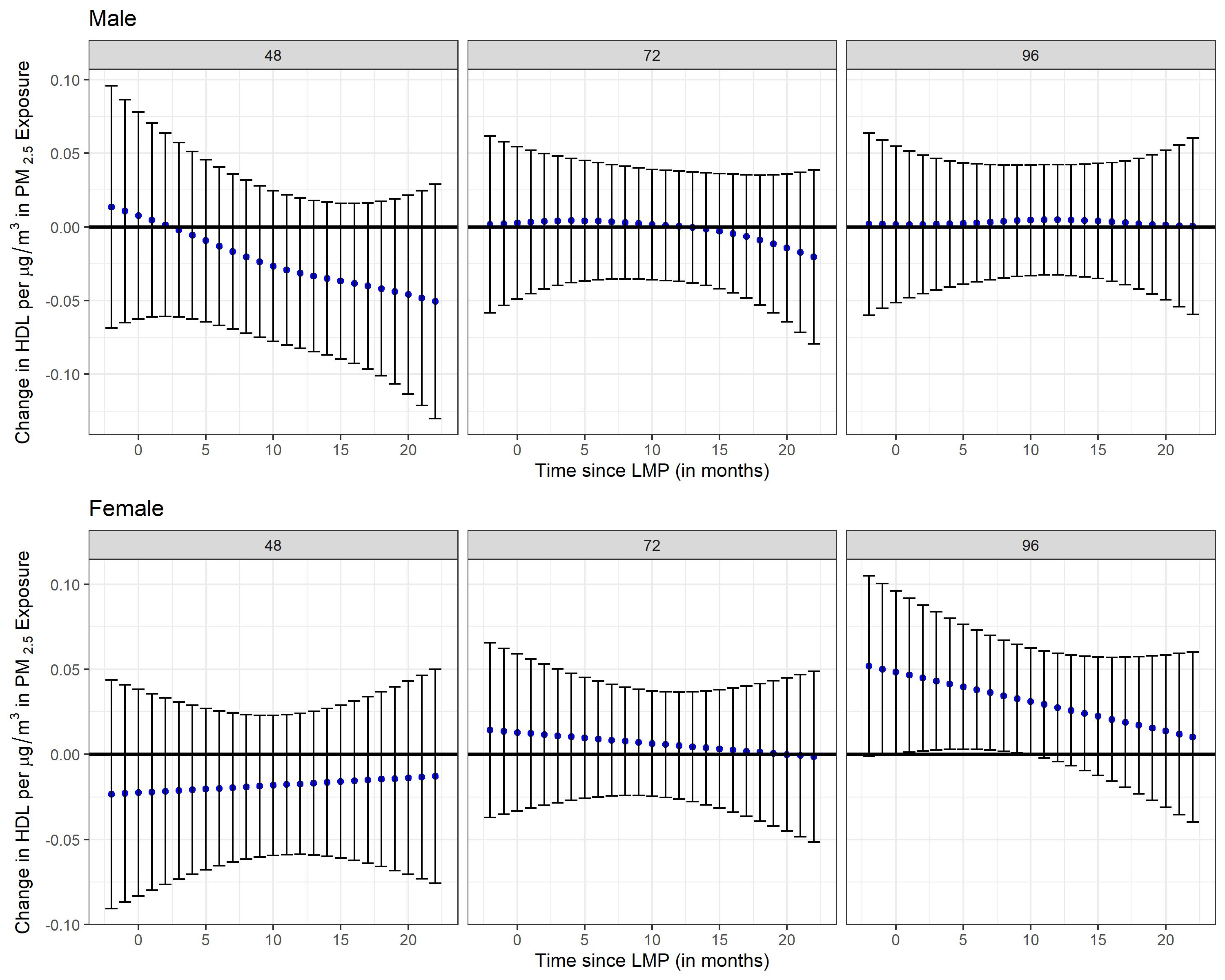

**Supplemental Figure 15.** Stratified analyses by fetal sex (pregnant mothers carrying either a male or female fetus) using distributed lag interaction models (DLIMs) with penalization and non-linear modification. Estimates of the association (blue dots) and 95% confidence interval (95% CI) (grey whiskers) between monthly PM_2.5_ exposure and low-density lipoprotein (LDL) at 48, 72, and 96 months after parturition. Negative and positive values on the x-axis indicate months before and after last menstrual period (LMP), respectively. Month 0 indicates the LMP month (month 0). Models were adjusted for age, pre-pregnancy BMI, socio-economic status, smoking during pregnancy, marital status, parity at enrollment, meteorological season, cardiometabolic medications, alcohol intake, multiple pregnancies throughout follow-up, stage, and follow-up time in months squared to allow for a nonlinear effect. Sample sizes for stratified analyses in pregnant mothers were n=267 for the male fetus carrier group and n=250 for the female fetus carrier group.

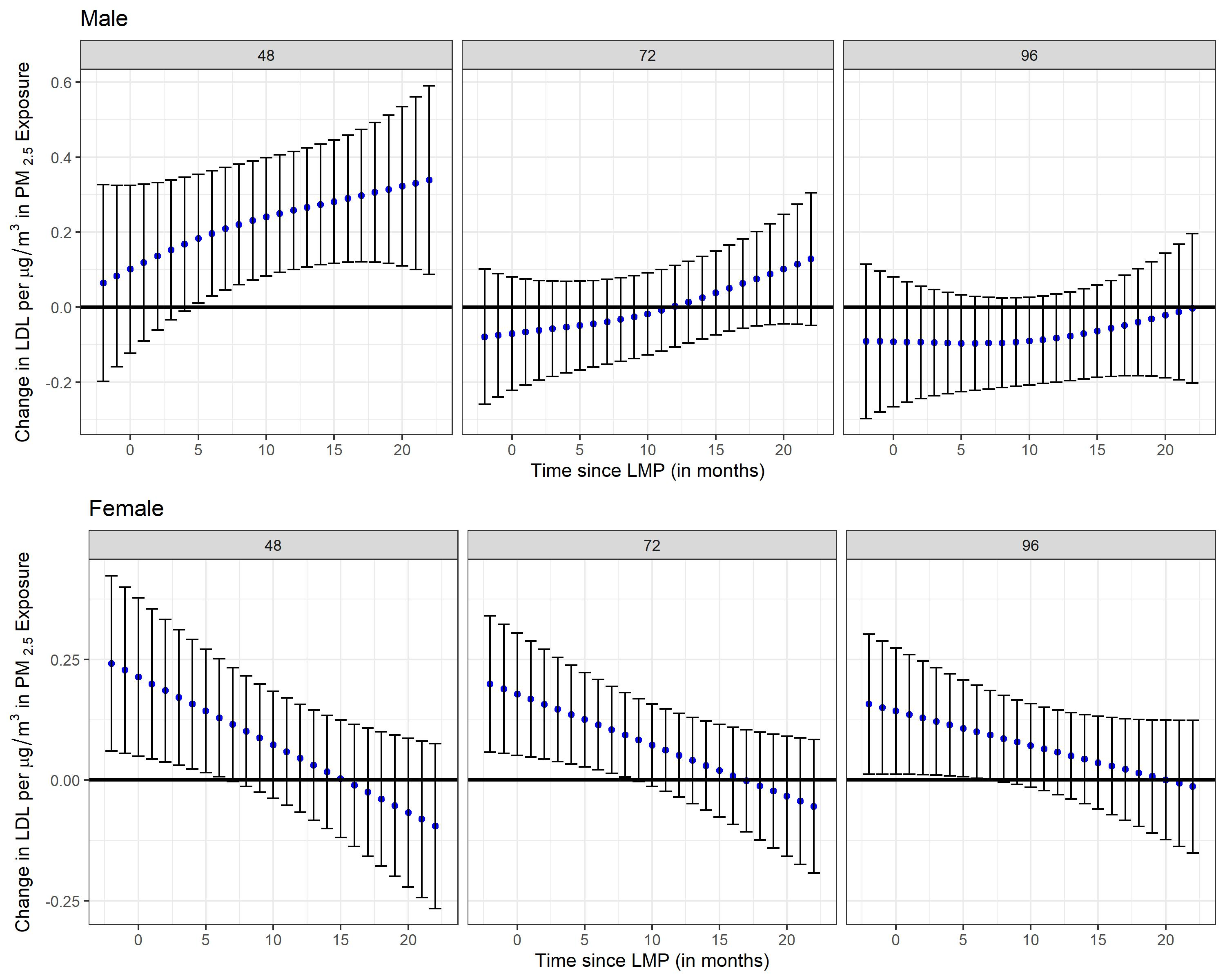

**Supplemental Figure 16.** Stratified analyses by folic acid (FA) supplementation during pregnancy (FA <600 mcg/day vs. FA ≥ 600 mcg/day) using distributed lag interaction models (DLIMs) with penalization and non-linear modification. Estimates of the association (blue dots) and 95% confidence interval (95% CI) (grey whiskers) between monthly PM_2.5_ exposure and Body Mass Index (BMI) at 48, 72, and 96 months after parturition. Negative and positive values on the x-axis indicate months before and after last menstrual period (LMP), respectively. Month 0 indicates the LMP month (month 0). Models were adjusted for age, pre-pregnancy BMI, socio-economic status, smoking during pregnancy, marital status, parity at enrollment, meteorological season, cardiometabolic medications, alcohol intake, multiple pregnancies throughout follow-up, stage, and follow-up time in months squared to allow for a nonlinear effect. Sample sizes for stratified analyses in pregnant mothers were n=338 for the low FA group (FA <600 mcg/day) and n=97 for the high FA group (FA ≥600 mcg/day).

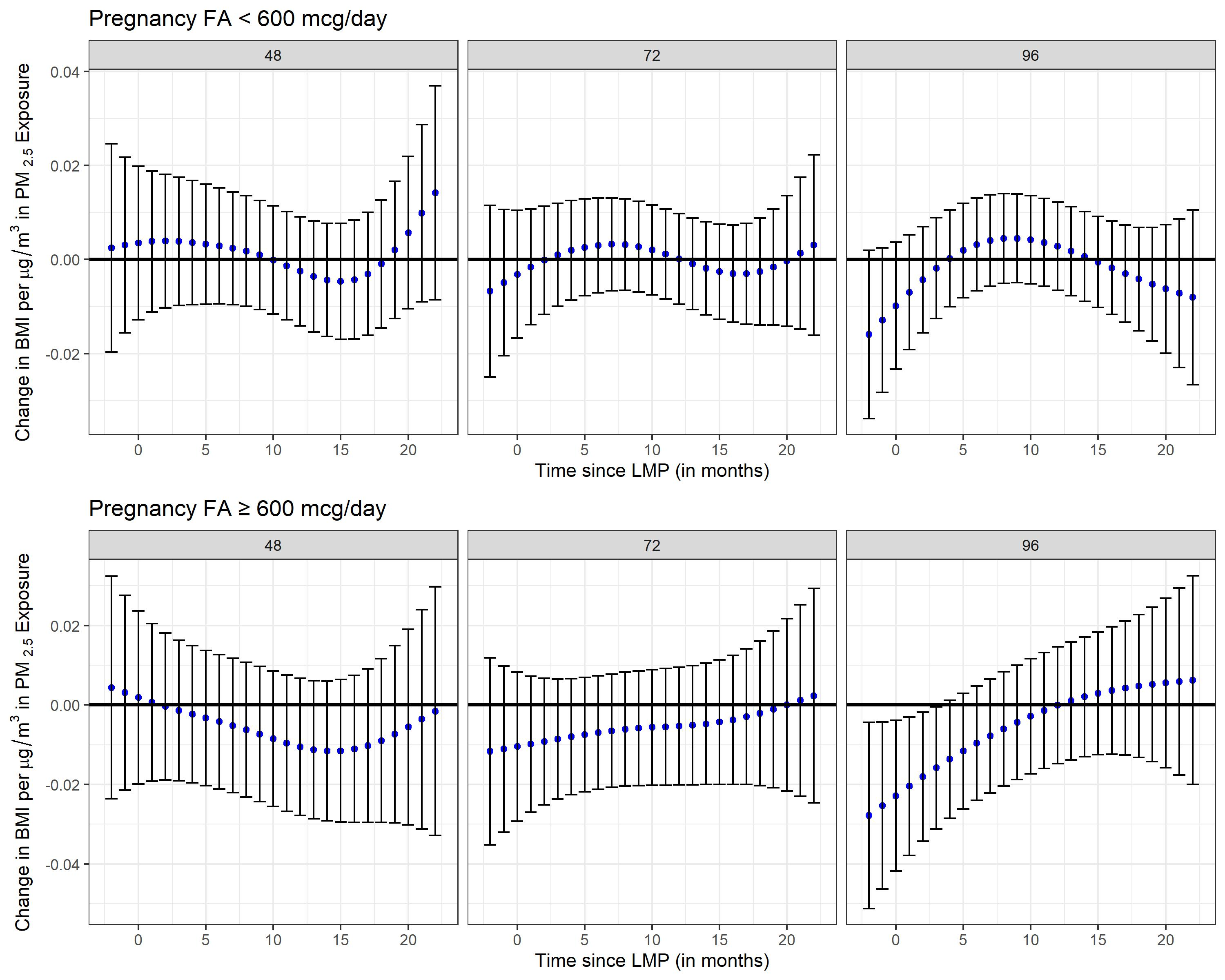

**Supplemental Figure 17.** Stratified analyses by folic acid (FA) supplementation during pregnancy (FA <600 mcg/day vs. FA ≥ 600 mcg/day) using distributed lag interaction models (DLIMs) with penalization and non-linear modification. Estimates of the association (blue dots) and 95% confidence interval (95% CI) (grey whiskers) between monthly PM_2.5_ exposure and Waist Circumference (WC) at 48, 72, and 96 months after parturition. Negative and positive values on the x-axis indicate months before and after last menstrual period (LMP), respectively. Month 0 indicates the LMP month (month 0). Models were adjusted for age, pre-pregnancy BMI, socio-economic status, smoking during pregnancy, marital status, parity at enrollment, meteorological season, cardiometabolic medications, alcohol intake, multiple pregnancies throughout follow-up, stage, and follow-up time in months squared to allow for a nonlinear effect. Sample sizes for stratified analyses in pregnant mothers were n=337 for the low FA group (FA <600 mcg/day) and n=97 for the high FA group (FA ≥600 mcg/day).

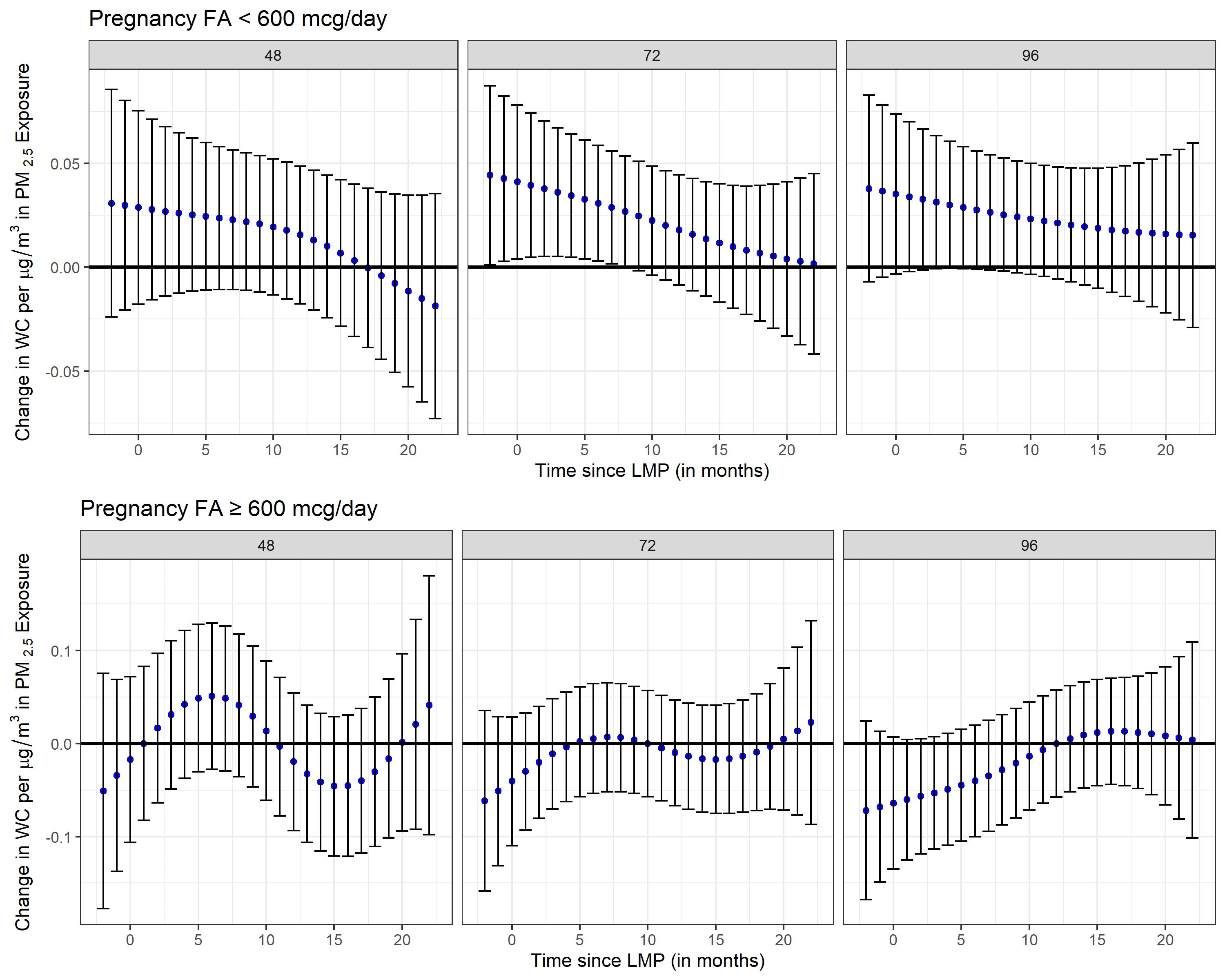

**Supplemental Figure 18.** Stratified analyses by folic acid (FA) supplementation during pregnancy (FA <600 mcg/day vs. FA ≥ 600 mcg/day) using distributed lag interaction models (DLIMs) with penalization and non-linear modification. Estimates of the association (blue dots) and 95% confidence interval (95% CI) (grey whiskers) between monthly PM_2.5_ exposure and body fat percentage at 48, 72, and 96 months after parturition. Negative and positive values on the x-axis indicate months before and after last menstrual period (LMP), respectively. Month 0 indicates the LMP month (month 0). Models were adjusted for age, pre-pregnancy BMI, socio-economic status, smoking during pregnancy, marital status, parity at enrollment, meteorological season, cardiometabolic medications, alcohol intake, multiple pregnancies throughout follow-up, stage, and follow-up time in months squared to allow for a nonlinear effect. Sample sizes for stratified analyses in pregnant mothers were n=337 for the low FA group (FA <600 mcg/day) and n=97 for the high FA group (FA ≥600 mcg/day).

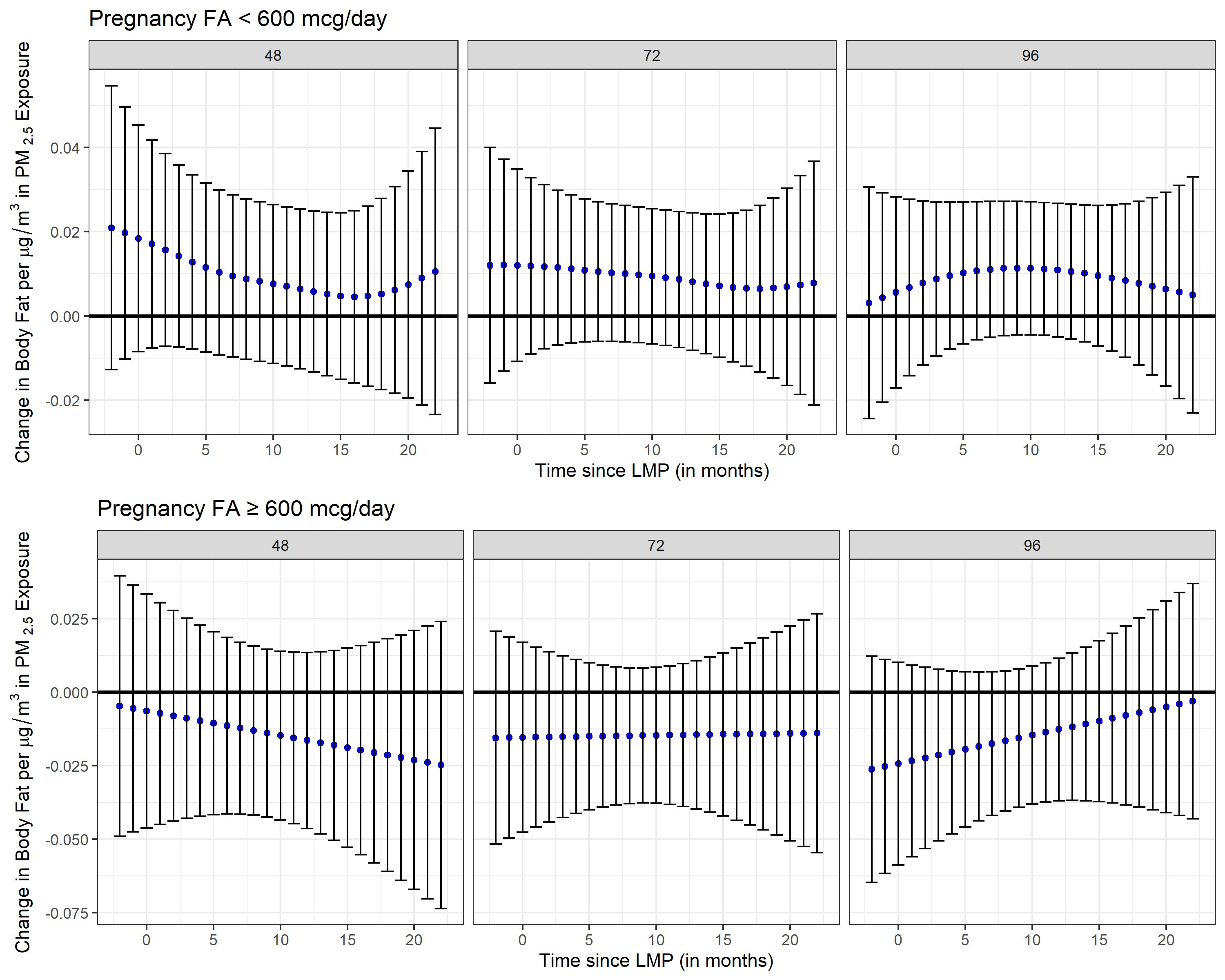

**Supplemental Figure 19.** Stratified analyses by folic acid (FA) supplementation during pregnancy (FA <600 mcg/day vs. FA ≥ 600 mcg/day) using distributed lag interaction models (DLIMs) with penalization and non-linear modification. Estimates of the association (blue dots) and 95% confidence interval (95% CI) (grey whiskers) between monthly PM_2.5_ exposure and total cholesterol at 48, 72, and 96 months after parturition. Negative and positive values on the x-axis indicate months before and after last menstrual period (LMP), respectively. Month 0 indicates the LMP month (month 0). Models were adjusted for age, pre-pregnancy BMI, socio-economic status, smoking during pregnancy, marital status, parity at enrollment, meteorological season, cardiometabolic medications, alcohol intake, multiple pregnancies throughout follow-up, stage, and follow-up time in months squared to allow for a nonlinear effect. Sample sizes for stratified analyses in pregnant mothers were n=339 for the low FA group (FA <600 mcg/day) and n=97 for the high FA group (FA ≥600 mcg/day).

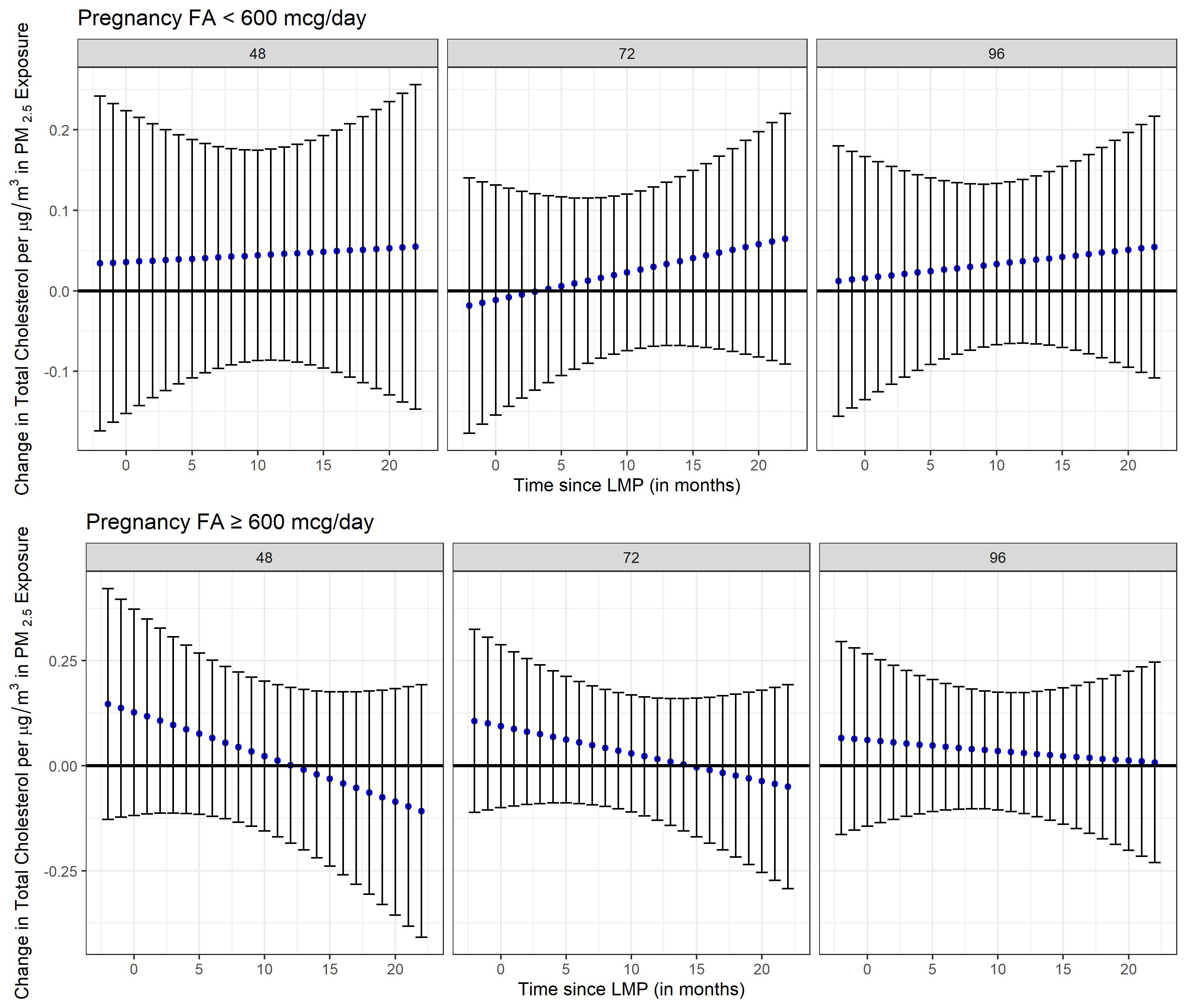

**Supplemental Figure 20.** Stratified analyses by folic acid (FA) supplementation during pregnancy (FA <600 mcg/day vs. FA ≥ 600 mcg/day) using distributed lag interaction models (DLIMs) with penalization and non-linear modification. Estimates of the association (blue dots) and 95% confidence interval (95% CI) (grey whiskers) between monthly PM_2.5_ exposure and high-density lipoprotein (HDL) at 48, 72, and 96 months after parturition. Negative and positive values on the x-axis indicate months before and after last menstrual period (LMP), respectively. Month 0 indicates the LMP month (month 0). Models were adjusted for age, pre-pregnancy BMI, socio-economic status, smoking during pregnancy, marital status, parity at enrollment, meteorological season, cardiometabolic medications, alcohol intake, multiple pregnancies throughout follow-up, stage, and follow-up time in months squared to allow for a nonlinear effect. Sample sizes for stratified analyses in pregnant mothers were n=339 for the low FA group (FA <600 mcg/day) and n=97 for the high FA group (FA ≥600 mcg/day).

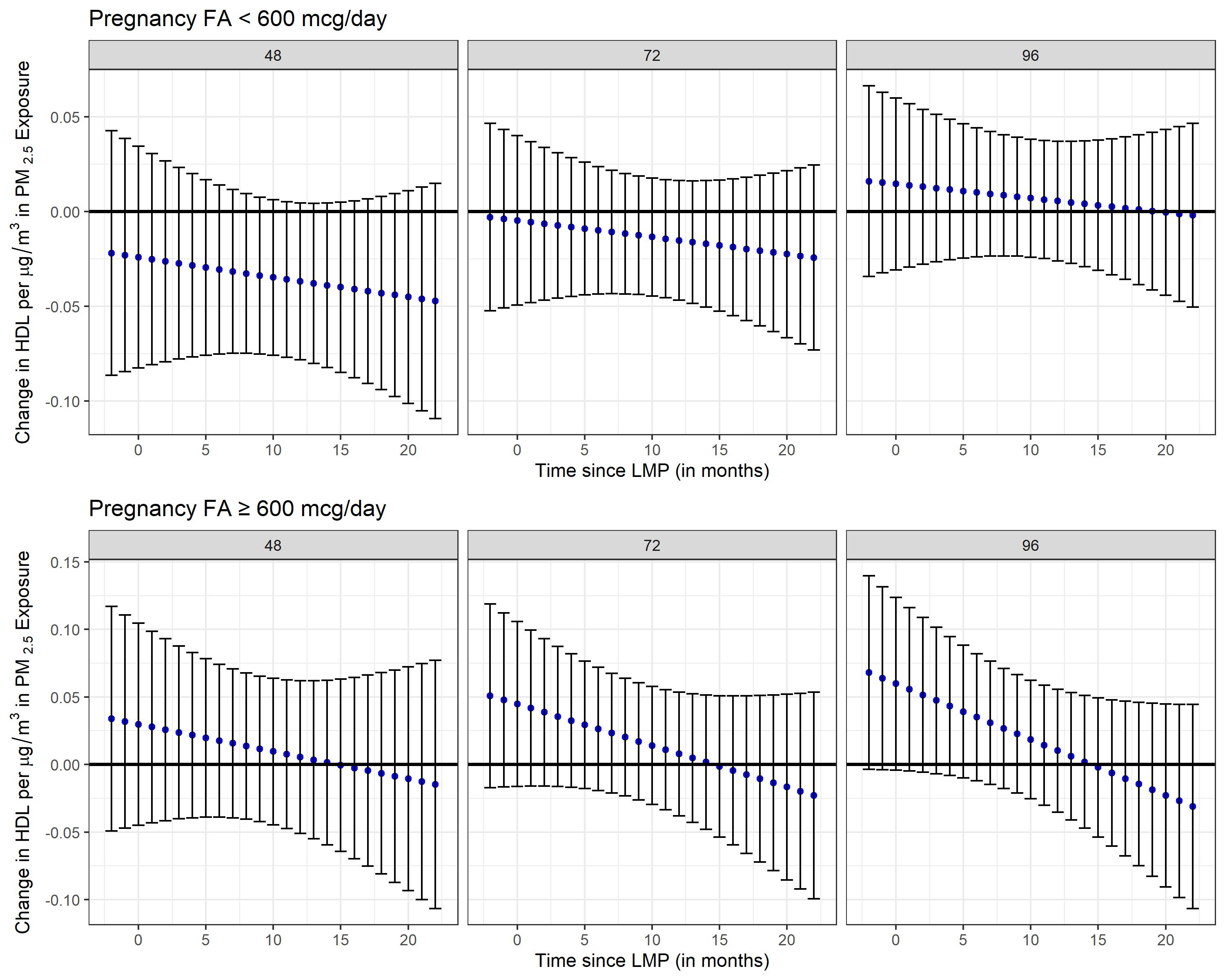

**Supplemental Figure 21.** Stratified analyses by folic acid (FA) supplementation during pregnancy (FA <600 mcg/day vs. FA ≥ 600 mcg/day) using distributed lag interaction models (DLIMs) with penalization and non-linear modification. Estimates of the association (blue dots) and 95% confidence interval (95% CI) (grey whiskers) between monthly PM_2.5_ exposure and low-density lipoprotein (LDL) at 48, 72, and 96 months after parturition. Negative and positive values on the x-axis indicate months before and after last menstrual period (LMP), respectively. Month 0 indicates the LMP month (month 0). Models were adjusted for age, pre-pregnancy BMI, socio-economic status, smoking during pregnancy, marital status, parity at enrollment, meteorological season, cardiometabolic medications, alcohol intake, multiple pregnancies throughout follow-up, stage, and follow-up time in months squared to allow for a nonlinear effect. Sample sizes for stratified analyses in pregnant mothers were n=339 for the low FA group (FA <600 mcg/day) and n=97 for the high FA group (FA ≥600 mcg/day).

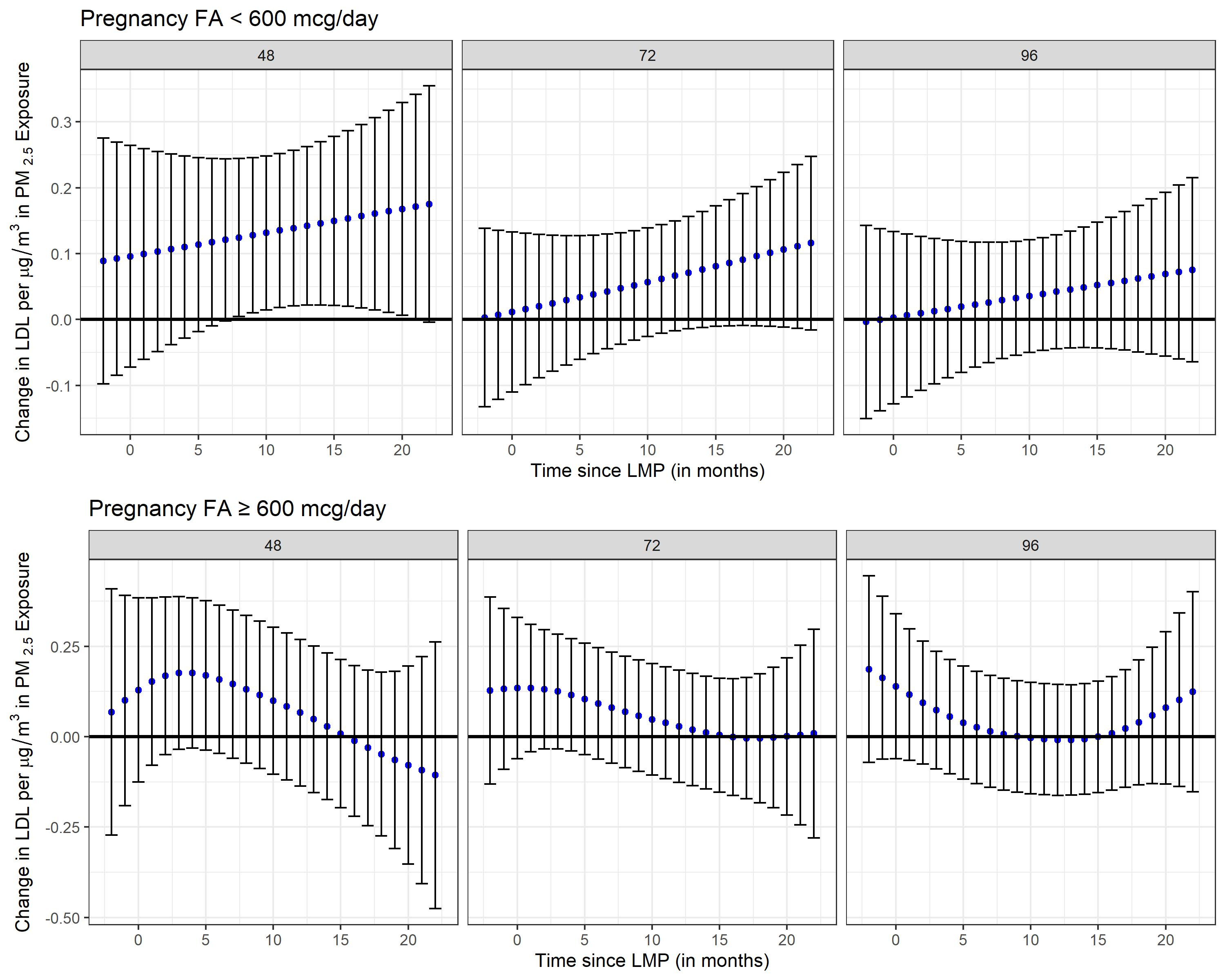
